# Host genome analysis of structural variations by Optical Genome Mapping provides clinically valuable insights into genes implicated in critical immune, viral infection, and viral replication pathways in patients with severe COVID-19

**DOI:** 10.1101/2021.01.05.21249190

**Authors:** Nikhil Shri Sahajpal, Chi-Yu Jill Lai, Alex Hastie, Ashis K Mondal, Siavash Raeisi Dehkordi, Cas van der Made, Olivier Fedrigo, Farooq Al-Ajli, Sawan Jalnapurkar, Rashmi Kanagal-Shamanna, Brynn Levy, Silviu-Alin Bacanu, Michael C Zody, Catherine A. Brownstein, Amyn M. Rojiani, Alan H. Beggs, Vineet Bafna, Alexander Hoischen, Erich D. Jarvis, Alka Chaubey, Ravindra Kolhe, the COVID19hostgenomesv consortium

## Abstract

**Background:** The varied clinical manifestations and outcomes in patients with SARS-CoV-2 infections implicate a role of host-genetics in the predisposition to disease severity. This is supported by evidence that is now emerging, where initial reports identify common risk factors and rare genetic variants associated with high risk for severe/ life-threatening COVID-19. Impressive global efforts have focused on either identifying common genetic factors utilizing short-read sequencing data in Genome-Wide Association Studies (GWAS) or whole-exome and genome studies to interrogate the human genome at the level of detecting single nucleotide variants (SNVs) and short indels. However, these studies lack the sensitivity to accurately detect several classes of variants, especially large structural variants (SVs) including copy number variants (CNVs), which account for a substantial proportion of variation among individuals. Thus, we investigated the host genomes of individuals with severe/life-threatening COVID-19 at the level of large SVs (500bp-Mb level) to identify events that might provide insight into the inter-individual clinical variability in clinical course and outcomes of COVID-19 patients.

**Methods:** Optical genome mapping using Bionano’s Saphyr® system was performed on thirty-seven severely ill COVID-19 patients admitted to intensive care units (ICU). To extract candidate SVs, three distinct analyses were undertaken. First, an unbiased whole-genome analysis of SVs was performed to identify rare/unique genic SVs in these patients that did not appear in population datasets to determine candidate loci as decisive predisposing factors associated with severe COVID-19. Second, common SVs with a population frequency filter was interrogated for possible association with severe COVID-19 based on literature surveys. Third, genome-wide SV enrichment in severely ill patients versus the general population was investigated by calculating odds ratios to identify top-ranked genes/loci. Candidate SVs were confirmed using qPCR and an independent bioinformatics tool (FaNDOM).

**Results:** Our patient-centric investigation identified 11 SVs involving 38 genes implicated in three key host-viral interaction pathways: (1) innate immunity and inflammatory response, (2) airway resistance to pathogens, and (3) viral replication, spread, and RNA editing. These included seven rare/unique SVs (not present in the control dataset), identified in 24.3% (9/37) of patients, impacting up to 31 genes, of which *STK26* and *DPP4* are the most promising candidates. A duplication partially overlapping *STK26* was corroborated with data showing upregulation of this gene in severely ill patients. Further, using a population frequency filter of less than 20% in the Bionano control dataset, four SVs involving seven genes were identified in 56.7% (21/37) of patients.

**Conclusion:** This study is the first to systematically assess and highlight SVs’ potential role in the pathogenesis of COVID-19 severity. The genes implicated here identify novel SVs, especially *STK26*, and extend previous reports involving innate immunity and type I interferon response in the pathogenesis of COVID-19. Our study also shows that optical genome mapping can be a powerful tool to identify large SVs impacting disease outcomes with split survival and add valuable genomic information to the existing sequencing-based technology databases to understand the inter-individual variability associated with SARS-CoV-2 infections and COVID-19 mortality.

## Introduction

The emergence of COVID-19 in the city of Wuhan, China in December 2019 led to an ongoing global pandemic. Since then, severe acute respiratory syndrome coronavirus 2 (SARS-CoV-2) has infected more than 80,721,623 individuals across the globe, with at least 1,763,690 COVID-19 related deaths (https://coronavirus.jhu.edu/map.html, last accessed December 27, 2020). The clinical manifestations of SARS-CoV-2 infected patients vary from asymptomatic or mild, including low-grade fever and flu-like symptoms, to severe symptoms, including acute respiratory distress syndrome (ARDS), pneumonia and death [**1-6**]. Clinical studies have associated advanced age, male gender, hypertension, diabetes, and other obesity-related diseases as risk factors predisposing to COVID-19 related severe illness [**7-9**]. However, variable clinical manifestations within these sub-sets and poor clinical outcome in individuals with no associated co-morbidities or risk factors clearly implicate the role of host genetics in the predisposition of SARS-CoV-2 infected individuals to disease severity [**10-11**]. A recent meta-analysis on host genetic factors in coronaviruses infection by LoPresti et al. [**12**] identified 1,832 relevant research publications with 105 of high significance. Of the 105 articles, seventy-five investigated human host genetic factors, identifying multiple significant loci, including 16 related to susceptibility (seven of which identified protective alleles) and 16 related to outcomes (three of which identified protective alleles). In addition, 30 articles investigated inter-species differences in disease susceptibility and pathogenesis by studying both human and non-human host genetic factors [**12**].

In principle, there have been two distinct approaches to investigate the host genome to identify genetic loci associated with disease susceptibility and severity in COVID-19 patients. The first approach utilizes relatively unbiased genome-wide association studies (GWAS) to understand risk factors at the population level, which provides insights into the pathophysiological mechanisms and biology, but rarely at an individual level. The second approach involves short-read whole exome or genome sequencing to identify rare variants in known genes whose biological functions suggest plausible models by which they may function as severe risk factors, such as those that contribute to inborn errors of immunity (IEIs) which remain unremarkable until infection, or completely novel IEIs/ primary immunodeficiencies (PIDs) that predispose to severe COVID-19. Using a GWAS approach, several groups have found that blood group A of the ABO groups is an independent risk factor for COVID-19 related disease susceptibility [**13, 14**]. In additional studies, chr3p21.31, chr12q24.13, chr19p13.2, chr19p13.3, and chr21q22.1 loci were associated with severe COVID-19, and blood group A was confirmed to be the risk factor for disease severity [**15, 16**]. Reports utilizing a targeted variant approach demonstrated the importance of rare/unique events that cause immunodeficiencies and predispose to severe COVID-19. For example, putative loss of function variants in *TLR7* in two different pairs of young and otherwise healthy brothers was associated with severe COVID-19 [**17**]. Further, the 13 genes in the TLR3/IRF3 pathway have been implicated in patients with severe COVID-19 [**18, 19**].

In a continued effort to understand the varied host response, the COVID-19 Host Genetics Initiative has been established to encourage data generation, sharing and meta-analysis of the GWAS statistics around the world [**20**]. Although these studies have identified certain genetic loci associated with disease severity, they remain limited to nucleotide variants, which explains only a small portion of the heritability of complex traits. Importantly, structural variants (SVs) are beyond the purview of these studies as a result of technical limitations, and they comprise a substantial proportion of genomic variation among individuals, which can drive evolutionary processes [**21-23**]. SVs are diverse, and include large copy-number changes, insertions, deletions, inversions, and translocations. SVs involve larger regions of an individual genome (500 bp and up) than small variants (up to several hundred basepairs). As most of these are not detected by routine short-read next generation sequencing and current analytical pipelines these categories of SVs remain undocumented with respect to their relationship with COVID-19 predisposition. According to the Human Gene Mutation Database (HGMD), more than 34% of all known disease-causing variation are larger than a single base-pair substitution, i.e. single nucleotide variation (SNVs) [**20**]. Several studies have demonstrated the importance of large SVs in the characterization of human immunity profiles [**24-26**]. Despite the host genome investigation initiatives across the globe to understand the genetic predisposition to disease severity in COVID-19, a substantial portion of the genome and variant classes remains intractable because of the technical limitations of applied genomic sequencing technologies [**26-28**].

To address this issue, we formed the COVID-19 Host Genome Structural Variation Consortium, and performed a preliminary study aimed at evaluating optical genome mapping to identify SVs in an unbiased fashion in severely ill patients with COVID-19. We hypothesize that structural variants in genes implicated in the viral response pathway(s), intractable by other genomic profiling techniques, may predispose some individuals to severe COVID-19.

## Materials and Methods

### Study Participants

A total of 37 severely ill COVID-19 patients defined as those admitted to the intensive care unit (ICU) who required mechanical ventilation or had a fraction of inspired oxygen (FiO2) of at least 60% or more, and with a confirmed SARS-CoV-2 RT-PCR test (from nasopharyngeal swabs or other biological fluids) were included in the study. The samples were collected under an approved HAC by the IRB Committee A (IRB REGISTRATION # 1597188-2), Augusta University, GA. Based on the IRB approval, all PHI was removed and all data was anonymized before accessing for the study.

### Optical Genome Mapping

Peripheral blood from critically ill COVID-19 patients was used to isolate ultra-high molecular weight (UHMW) DNA following the manufacturer’s protocols (Bionano Genomics, San Diego, USA). Briefly, a frozen blood aliquot (650μl) was thawed and cells were counted using HemoCue (HemoCue Holding AB, Ängelholm, Sweden). Subsequently, a blood aliquot comprising approximately 1.5 million nucleated white blood cells was centrifuged and digested with Proteinase K. DNA was precipitated using isopropanol and washed using buffers (buffer A and B), while the DNA remained adhered to the nanobind magnetic disk. The UHMW bound DNA was resuspended in elution buffer and quantified using Qubit dsDNA assay kits (ThermoFisher Scientific, San Francisco, USA).

DNA labeling was performed at the specific 6-base sequence motifs following manufacturer’s protocols (Bionano Genomics, USA) in which Direct Labeling Enzyme 1 (DLE-1) reactions to a known repeated sequence in the genome were carried out using 750 ng of purified high molecular weight DNA. Labeled DNA was loaded on to flow cells of Saphyr chips for optical imaging. The fluorescently labeled DNA molecules were imaged after the labelled DNA molecules were electrophoretically linearized, using the Bionano Genomics Saphyr® platform. Effective genome coverage of approximately 100X was achieved for tested samples after evaluating the molecule quality metrics. The quality control metric for each sample achieved the recommended molecule map rates of greater than 70% and molecule N50 values greater than 250kb.

Genome analyses were performed using Bionano Access (v.1.5)/Bionano Solve (v.3.5) software, a *de novo* assembly analysis was performed on all the samples to assess and interrogate all germ line SVs. Briefly, molecules of a given sample dataset were first de novo assembled into consensus genome maps, the genome maps were aligned to the hg19 reference human genome assembly. SVs were identified where *de novo* assemblies differed from the hg19 reference genome, insertion, duplications, deletions, inversions, and translocations could be called based on this alignment. SVs generated by the de novo assembly pipeline were then annotated with known canonical gene sets extracted from the reference genome assembly and compared to a control dataset to estimate the population frequency of SVs.

### Data analysis

To identify SVs associated with disease severity, three distinct analyses were undertaken. **In the first analysis**, rare/unique SVs were investigated to determine candidate gene/loci as potentially strong predisposing factors associated with severe COVID-19. An unbiased whole genome analysis of SVs was performed to identify unique/rare genic SVs in these patients that did not appear in the population dataset. Additionally, only the SVs disrupting the coding region(s) of the gene(s) were selected and reviewed for relevance in the COVID-19 infection. **In the second analysis**, common SVs were interrogated for possible association with severe COVID-19 based on a literature survey. A thorough literature review was performed with the combination of the following key words that included “SARS”, “Viral infection”, “COVID-19”, “host genetics”, “host susceptibility”, “immune response”, and “immune genes”. The literature included electronic, ahead-of-print, and pre-print articles which investigated DNA-based genetic variations, proteomics, transcriptomic, and bio-informatics modelling to identify candidate pathways/ genes. A gene list comprising of 881 genes was manually curated after searching the articles (last accessed November 15, 2020) for relevance pertaining to a general method, genes, variants, statistical results, associations, and p-value (**Supplementary file 1**). The SVs overlapping the genes of interest in severely ill patients were compared with a population dataset [Bionano Genomics (Bionano) controls] with **<20%** frequency filter. The SVs were further interrogated against the African American ethnicity in the Bionano control dataset [**29-31**]. Subsequently, only variants disrupting the coding regions of genes of interest were selected for further analysis. **In the third analysis**, genome-wide SV enrichment in severely ill patients versus a general population cohort (Bionano controls comprising 267 individuals, including 150 assayed with DLE-1 enzyme) was investigated by calculating odds ratio to identify top ranked genes/loci, with the following filtering criteria that includes, SVs observed in <20% of Bionano controls. The odds ratio of a gene was considered equal to the proportion of total severely ill patients having at least a SV on the gene, divided by proportion of total Bionano controls (150 individuals labeled with DLE-1 enzyme; **Figure 1**).

**Figure 1:**
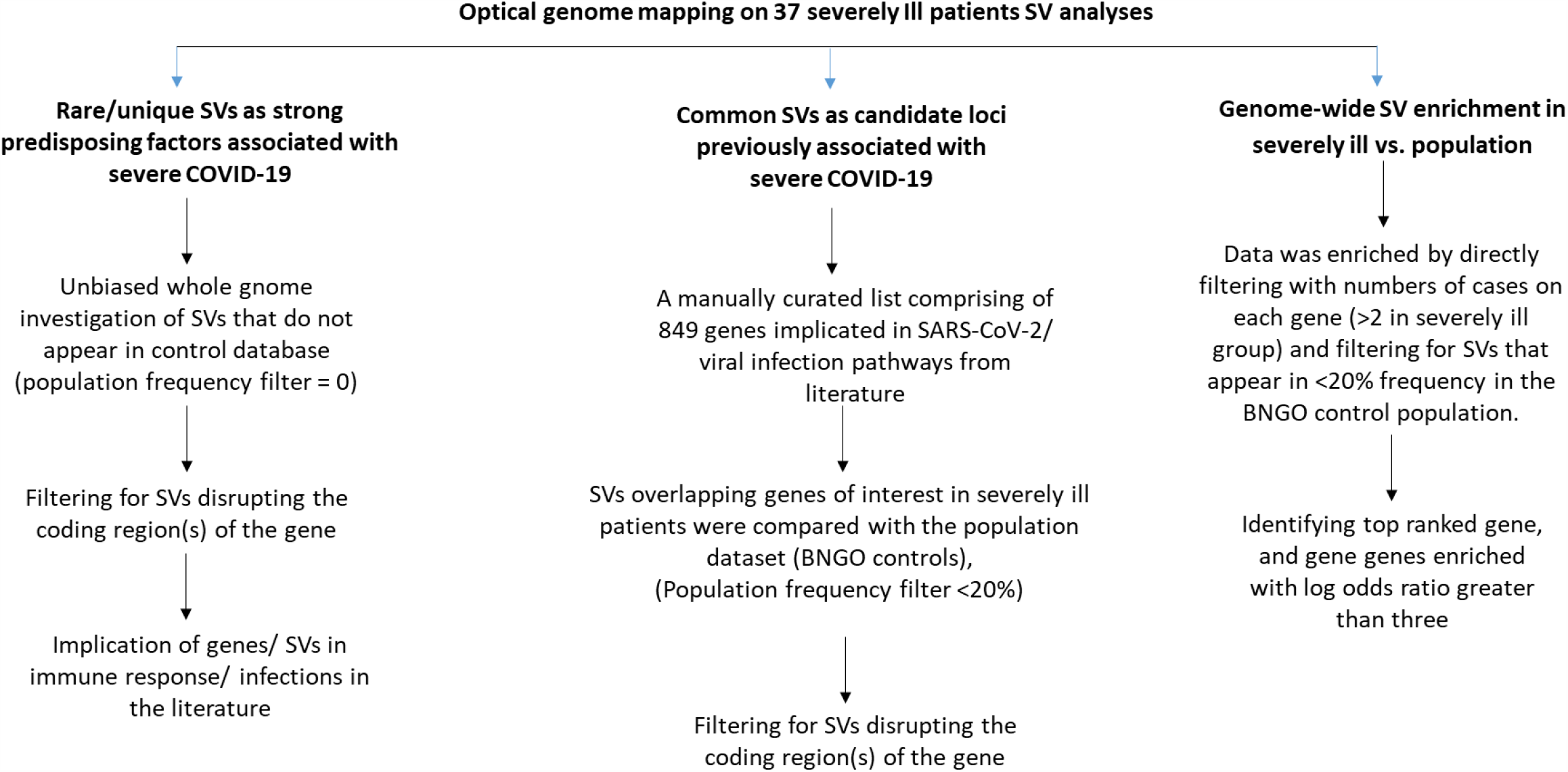
Workflow diagram for analysis of COVID-19 patient SVs.

### Quantitative PCR (qPCR) confirmation of OGM SVs

Quantitative PCR (qPCR) was used to confirm the selected SVs (copy number gains and losses). Relative quantitation of genomic dosage was determined using the QuantStudio 3 real time cycler (Thermo Fisher Scientific, San Francisco, USA) and calculated by the relative threshold cycle (ddCt) method [**32**]. PCR amplicons were generated in triplicate for each individual and four controls. Genomic dosage for the selected genes (*EDARADD, APOBEC3B, ZNF443, CCL4L2*) was determined by SYBR green incorporation using RNAseP as the reference (BioRad, USA). Three primer pairs were designed for each SV using the Primer 3.0 software (List of primer sequences and targets is available in **Supplementary file 2**). Relative genomic dosage was calculated as 2^-ΔΔCT^ where ΔCT=(mean Ct_Target_)-(mean Ct_Reference_) and ΔΔCT= ΔCT_patient_-ΔCT_control_.

### Independent assessment of Saphyr SVs by FaNDOM

To provide an independent assessment of the SVs called by the Bionano Solve pipelines developed by Bionano Genomics, we used a novel method, FaNDOM (Fast Nested Distance-based seeding of Optical Maps) (https://github.com/jluebeck/FaNDOM). FaNDOM maps optical map fragments to an *in silico* digested reference genome, relying on a fast filtering strategy to provide an order of magnitude speedup while still maintaining high sensitivity in excess of 95% for concordant reads, and 79% for SVs on the benchmark NA12878 human genome. FaNDOM was used on the COVID-19 patient data to call breakpoints (two distinct reference positions that are adjacent in the donor), multi-chromosomal translocation and insertion events. SVs were called on breakpoints based on the orientation of the fragments mapping to the breakpoint, with a (+,+) orientation suggestive of a deletion, (-,-) orientation suggestive of a tandem duplication, and opposing orientation suggesting an inversion or translocation. The SVs called by FaNDOM were used to independently confirm SVs called by Bionano Access Software.

### Expression Analysis

Total RNA was isolated from peripheral blood of 11 asymptomatic patients [(3 Male, 8 female) age range 25-59 years [43.2 (mean) ± 11.3 (Stdev)] and 12 severely ill COVID-19 patients [(6 Male and 6 Female) age range 18-81 years [(61.8 (mean) ± 15.4 (Stdev)], using mRNeasy mini kit (QIAGEN, Germany). Peripheral blood was collected from the asymptomatic controls during active infection (positive for SARS-CoV-2 by RT-PCR at time of blood collection), and during ICU treatment for the severely ill patients. The quantity of the total RNA from the samples was determined by ultraviolet spectrophotometer (Nanodrop, Thermo Fisher Scientific, and Pittsburgh, PA). Total RNA (500 ng) was reverse transcribed using the iScript cDNA synthesis kit (170–8891) from BioRad laboratories (Hercules, CA). Quantitative real-time PCR (q-RTPCR) was performed using gene-specific primers, and a SYBR Green assay on the QuantStudio 3 system (Thermo Fisher Scientific, CA). The specific products were confirmed by SYBR green single melt curve analysis. The results were normalized to the expression of the 18S rRNA hsousekeeping gene and the relative fold change was calculated using delta-delta Ct method.

## Results

### Patient Characteristics

During the period from March 30 to August 13, 2020, 37 severely ill patients were identified for this study. The criteria for inclusion included a confirmed SARS-CoV-2 RT-PCR test (from nasopharyngeal swabs or other biological fluids), and a requirement for mechanical ventilation or a fraction of inspired oxygen (FiO2) of at least 60% or more. The demographics and clinical characteristics of the patients (**Table 1**) were a mean age of 61.6 ±12.4 SD (range 19-83), 49% (18/37) female, and 70.2% (26/37) African American, 27% (10/37) Caucasian, and one Hispanic, which is similar to the demographics of COVID-19 patients in the State of Georgia, USA [**33**]. Chronic preexisting medical conditions in these patients included diabetes (43.2%), hypertension (59%), chronic kidney disease (24%), and asthma (5.4%). Four patients had no known co-morbidities at the time of admission to the ICU. On ICU admission, 30 patients needed mechanical ventilation, with mean intubation duration of 12 ± 5.5 days, of which, 35% (13/37) were intubated prior to admission (PTA), and 16% (6/37) were ventilated in prone-position. Of the 37 patients, 25 recovered and 12 died.

**Table 1.**
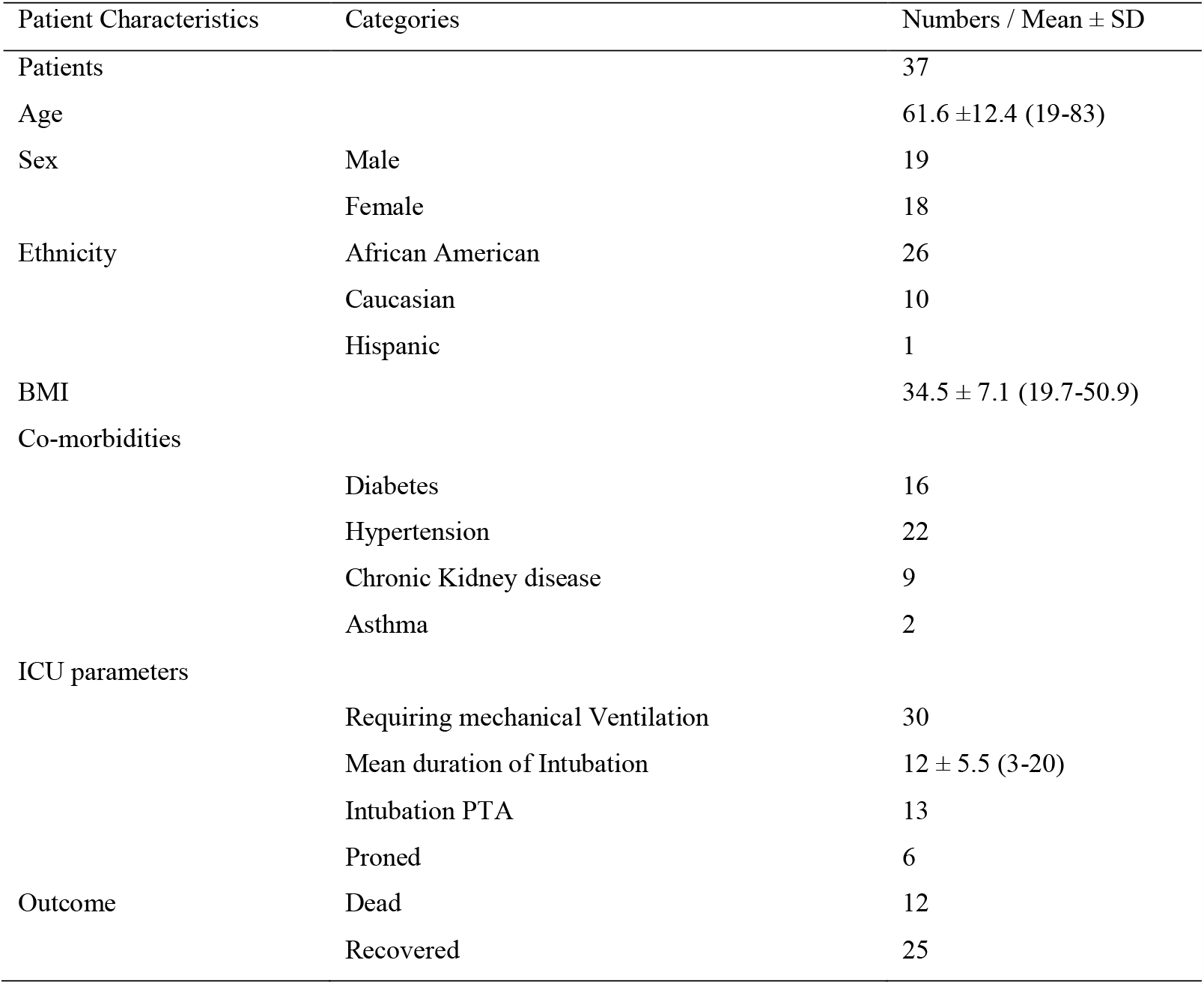
Clinical characteristics of patients included in the study.

### Rare/unique SVs as strong predisposing factors associated with severe COVID-19

An unbiased whole genome mapping investigation of the severely ill patients was performed using a filter for SVs that did not appear in the Bionano controls. The complete list of rare/unique SVs is provided a **Supplementary file 3**. Private SVs disrupting the gene coding regions led to the identification of seven rare/unique SVs, possibly impacting 31 genes, identified in 24.3% (9/37) patients. These included: a ∼162.2 kb duplication of chr 1 (chr1:236604233-236766495) partially disrupting the *EDARADD* (exon 5 and 6; NM_145861.3) and *HEATR1* (exons 3-45; NM_018072.6) genes in two patients (cases 22 and 26); a ∼24.06 kb heterozygous deletion of chr 2 (chr2:162887379-162911439) partially deleting the *DPP4* gene (exons 3 and 4; NM_001935.4) in one patient (case 38); a ∼146.8 kb heterozygous deletion of chr 16 (chr16:67308198-67455019) that contained six genes (*PLEKHG4, KCTD19, LRRC36, TPPP3, U1, ZDHHC1*) was identified in one patient (case 39); a ∼ 28.4 kb duplication of chr 17 (chr17:39662399-39690882) including two genes (*KRT15* and *KRT19*) in one patient (case 2); a ∼833 kb **tandem** duplication of chr 17 (chr17:71844581-72678517) including 15 genes (*RPL38, MGC16275, TTYH2, Z49982, DNAI2, CD300E, CD300LD, CD300C, CD300LB, CD300A, GPRC5C, GPR142, BTBD17, KIF19)*, and partially disrupting *RAB37* (exon 1; NM_175738.5) in one patient (case 19); a ∼39.8 kb duplication of chr 19 (chr19:12512276-12552113) including the entire *ZNF443* gene (NM_005815.5) in two patients (cases 13 and 18); and a ∼ 534.9 kb tandem duplication of chr X (chrX: 130629618-131164603) including four genes (*OR13H1, LOC286467, 5S_rRNA, STK26*) in one patient (case 44) (**Table 2, Figure 2a-g)**. All these SVs were confirmed by qPCR (**Supplementary file 4**).

**Table 2.**
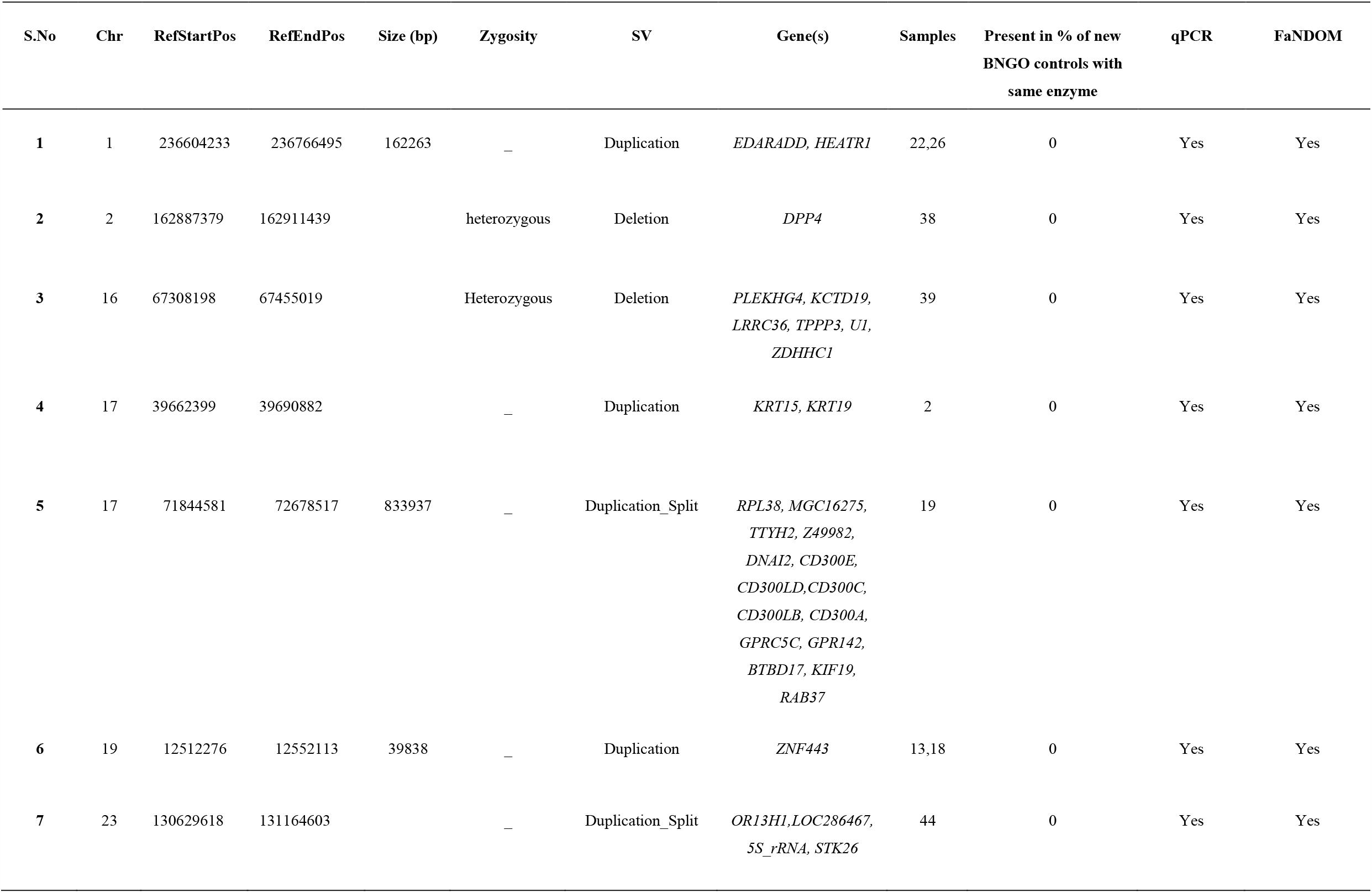
List of unique structural variants overlapping coding region(s) of gene(s) identified in patients with severe COVID-19.

**Figure 2a:**
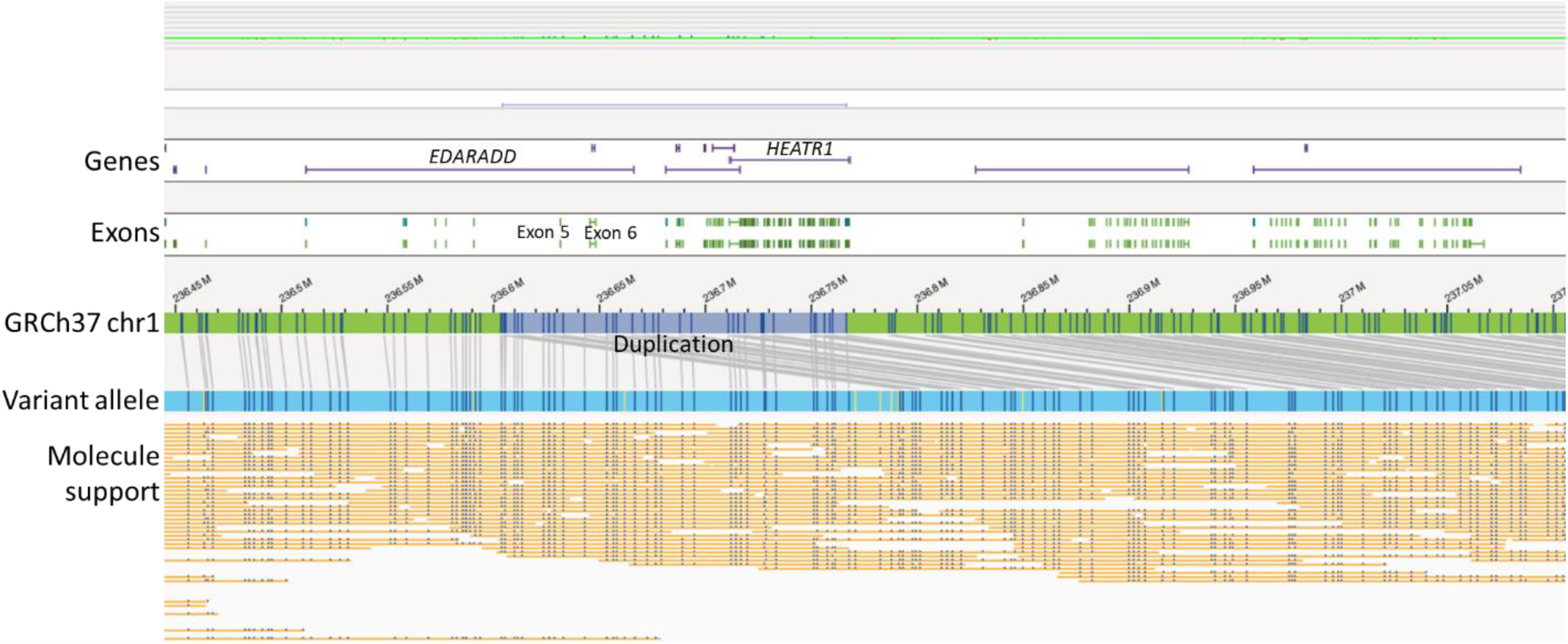
A ∼162.2 kb duplication of chr 1 (chr1:236604233-236766495) partially disrupting the *EDARADD* (exon 5 and 6; NM_145861.3) and *HEATR1* (exons 3-45; NM_018072.6) genes in two patients (cases 22 and 26).s

**Figure 2b:**
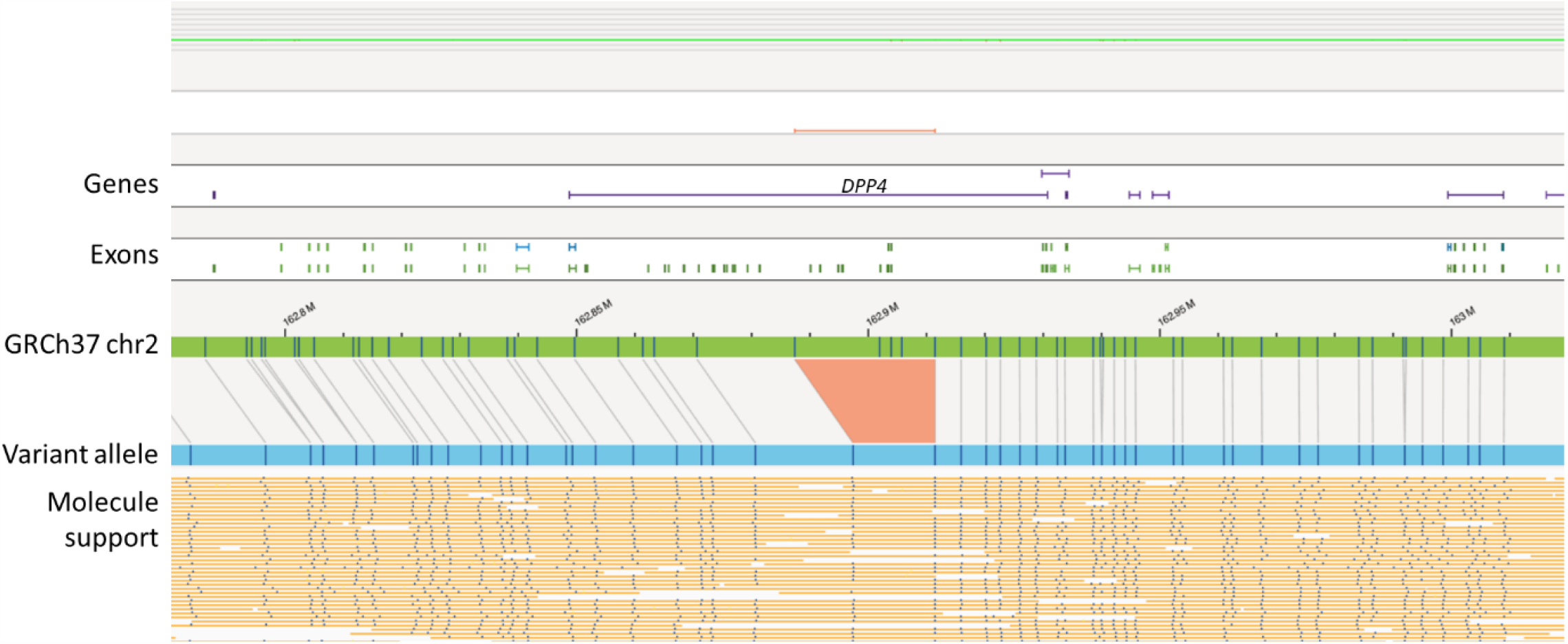
**A** ∼24.06 kb heterozygous deletion of chr 2 (chr2:162887379-162911439) partially deleting the *DPP4* gene (exons 3 and 4; NM_001935.4) in one patient (case 38).

**Figure 2c:**
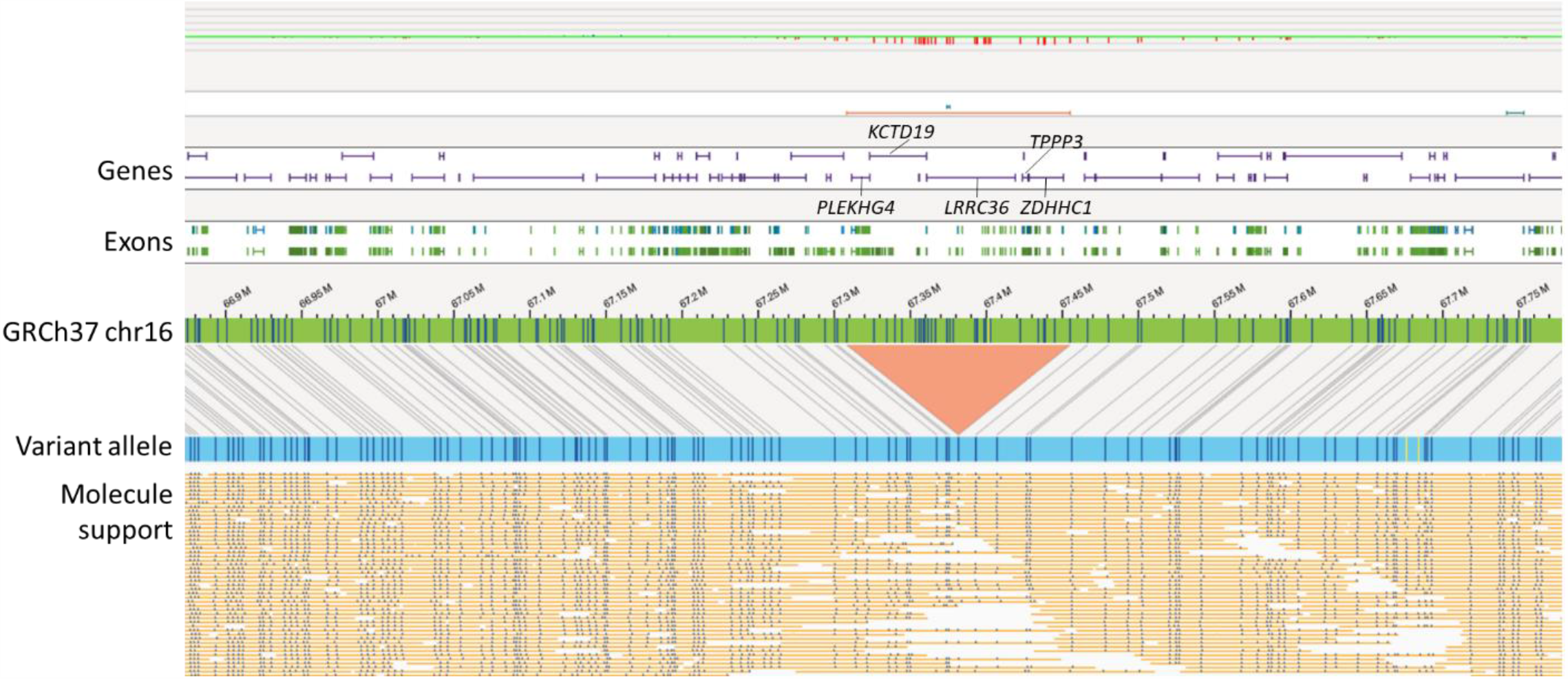
A ∼146.8 kb heterozygous deletion of chr 16 (chr16:67308198-67455019) that contained six genes (*PLEKHG4, KCTD19, LRRC36, TPPP3, U1, ZDHHC1*) was identified in one patient (case 39)

**Figure 2d:**
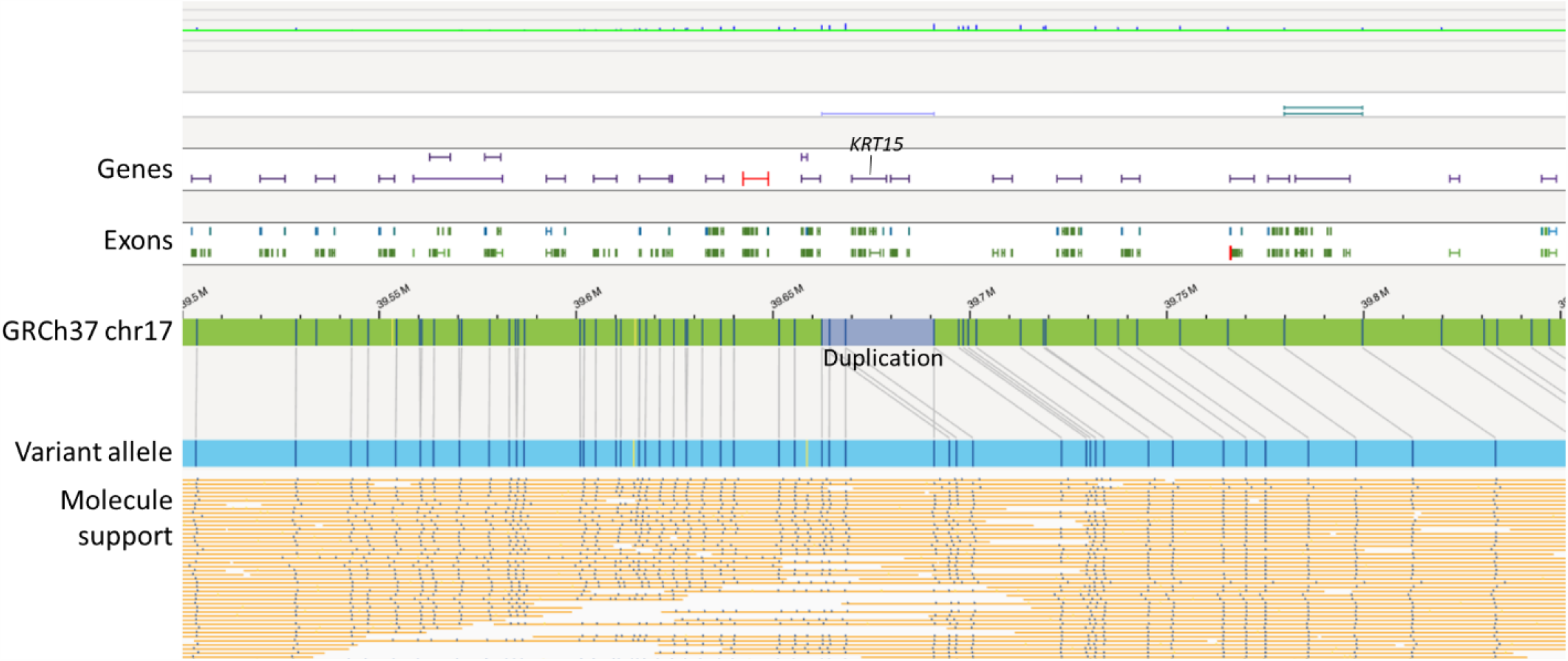
**A** ∼ 28.4 kb duplication of chr 17 (chr17:39662399-39690882) including two genes (*KRT15* and *KRT19*) in one patient (case 2).

**Figure 2e:**
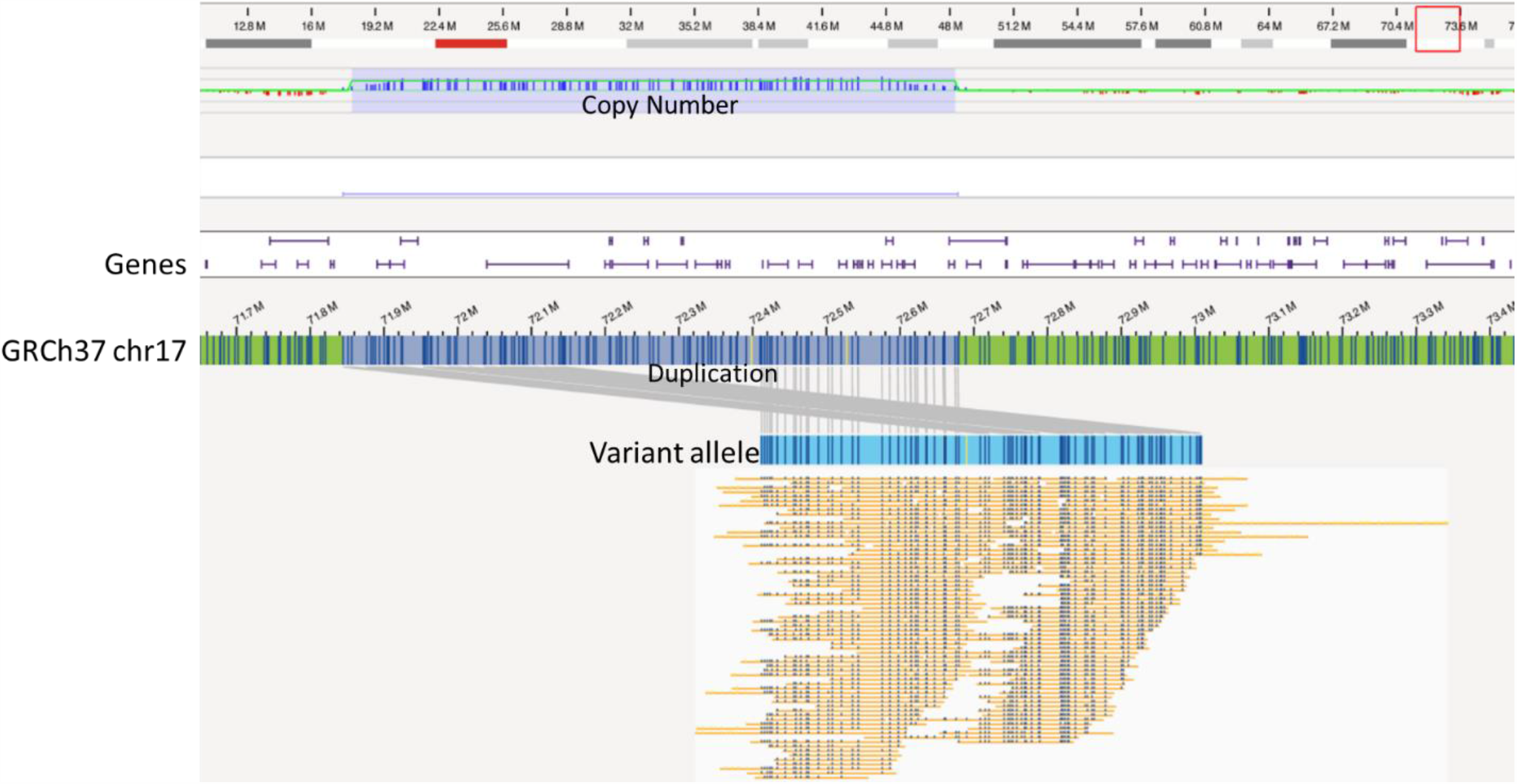
**A** ∼833 kb **tandem** duplication of chr 17 (chr17:71844581-72678517) including 15 genes (*RPL38, MGC16275, TTYH2, Z49982, DNAI2, CD300E, CD300LD, CD300C, CD300LB, CD300A, GPRC5C, GPR142, BTBD17, KIF19)*, and partially disrupting *RAB37* (exon 1; NM_175738.5) in one patient (case 19).

**Figure 2f:**
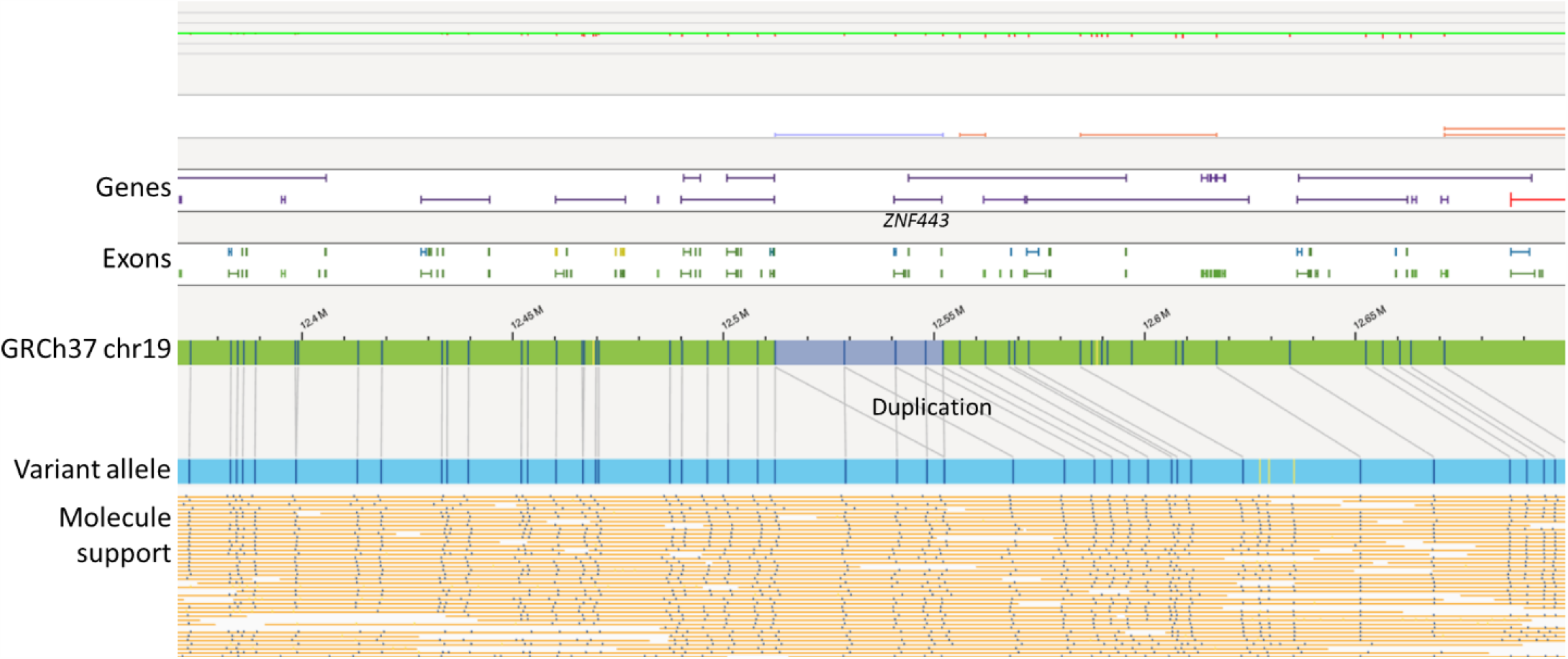
A ∼39.8 kb duplication of chr 19 (chr19:12512276-12552113) including the entire *ZNF443* gene (NM_005815.5) in two patients (cases 13 and 18).

**Figure 2g:**
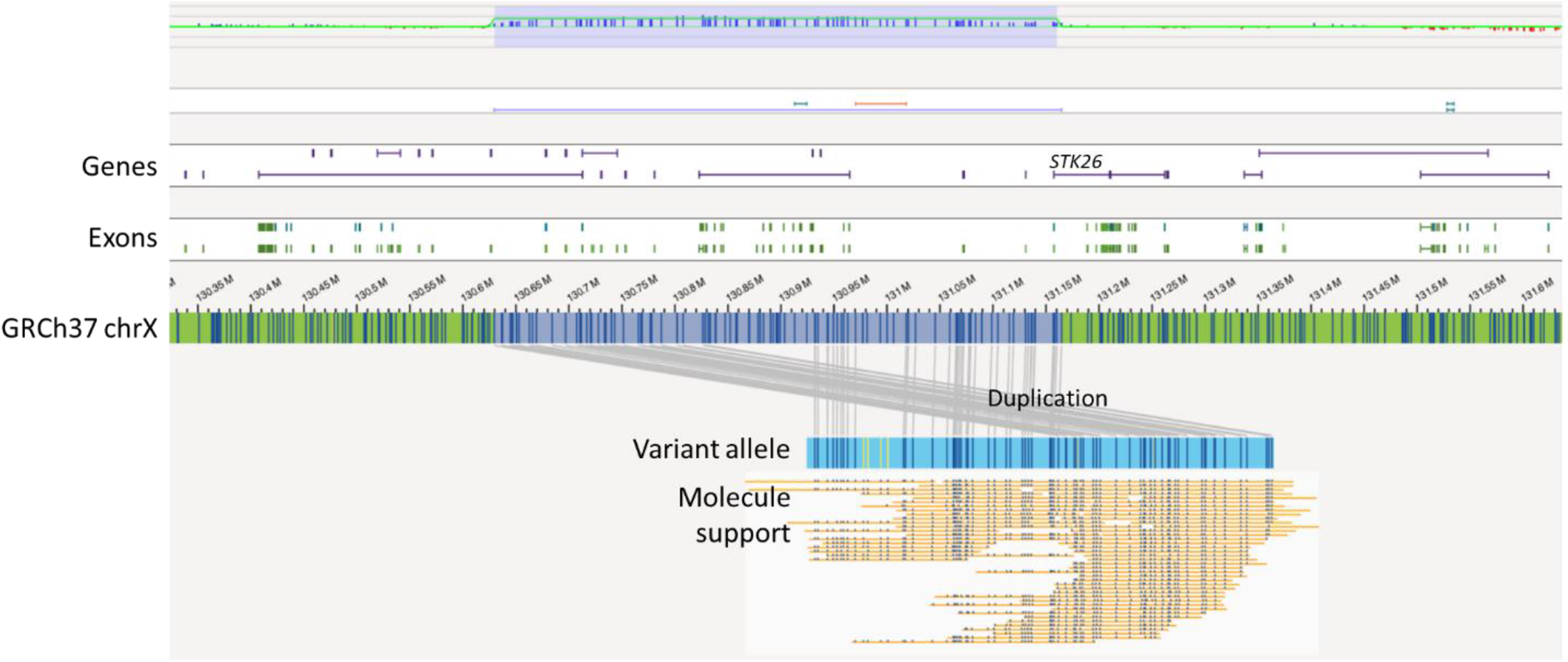
A ∼ 534.9 kb tandem duplication of chr X (chrX: 130629618-131164603) including four genes (*OR13H1, LOC286467, 5S_rRNA, STK26*) in one patient (case 44).

### Common SVs in candidate loci previously associated with severe COVID-19

We next searched for common SVs that overlap with previously suggested candidate loci/genes implicated in COVID-19. This literature-based approach involved investigating SVs overlapping coding regions of the 881 genes identified in literature survey. These SVs were further filtered by considering only those at frequency of less than 20% in Bionano control data sets (267 individuals). This filtering resulted in identification of four SVs impacting seven genes in 56.7% (21/37) severely ill patients. Based on the OGM data and the minimum genomic coordinates identified by the Bionano Access software, the four SVs identified include: a ∼248.1 kb heterozygous inversion of chr 3 (chr3:195725611-195477445) disrupting the *MUC4* gene (exons 1-23; NM_018406.7) in five patients of African American (AA) ethnicity (cases-2, 3, 8, 26, 29); A ∼29.8 kb duplication of chr 17 (chr17:34437711-34571682) including 3 genes (*TBC1D3B, CCL3L1, CCL4L2*) in ten patient (cases 12, 20, 28, 29, 6, 8, 13, 19, 21, 22); a ∼28.9 kb heterozygous deletion of chr 22 (chr22:39347202-39391164) disrupting 2 genes (*APOBEC3B* and *APOBEC3A*) in three patients; and a ∼1.5 kb insertion in chr 21 (chr21:42784603-42793518) overlapping *MX1* gene (exon 1; NM_001144925.2) in 11 patient (cases 13, 17, 18, 20, 21, 24, 25, 28, 4, 8, 38) (**Table 3, Figure 3a-d**). All SVs, except the *MUC4* inversions and *MX1* insertion were confirmed by qPCR (**Supplementary file 5**).

**Table 3.**
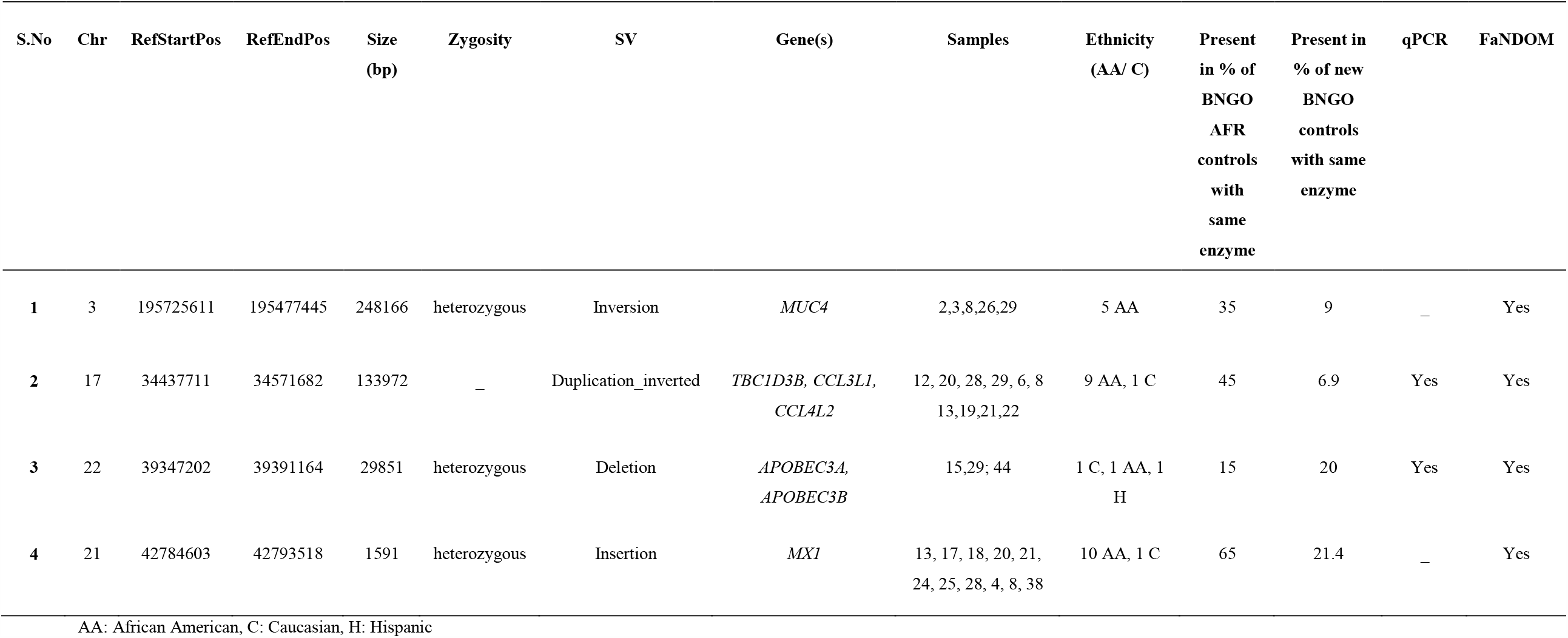
List of structural variants overlapping coding region(s) of gene(s) previously associated with COVID-19/ viral infections/ immune response identified in severe COVID-19 patients.

**Figure 3a:**
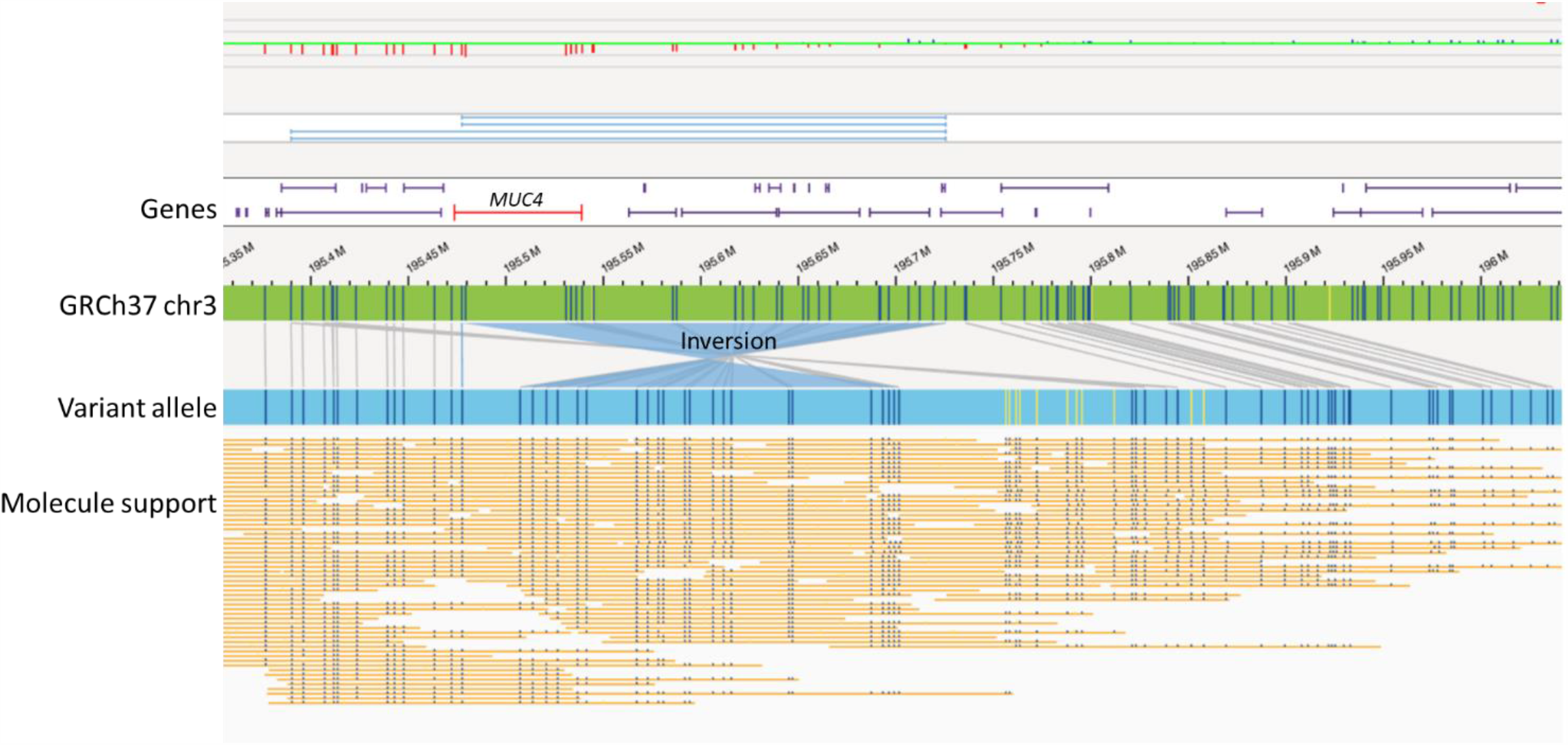
A ∼248.1 kb heterozygous inversion of chr 3 (chr3:195725611-195477445) disrupting the *MUC4* gene (exons 1-23; NM_018406.7) in five patients of African American (AA) ethnicity (cases-2, 3, 8, 26, 29).

**Figure 3b:**
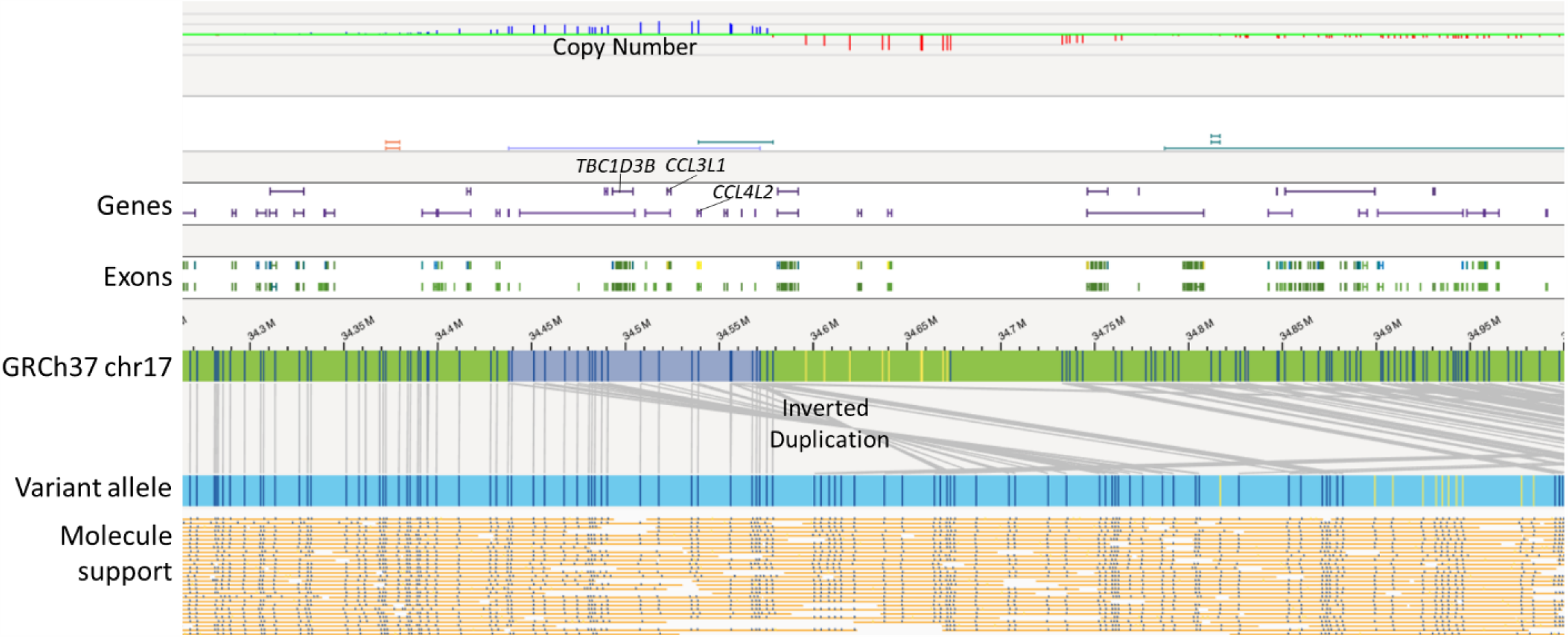
A ∼29.8 kb duplication of chr 17 (chr17:34437711-34571682) including 3 genes (*TBC1D3B, CCL3L1, CCL4L2*) in ten patient (cases 12, 20, 28, 29, 6, 8, 13, 19, 21, 22).

**Figure 3c:**
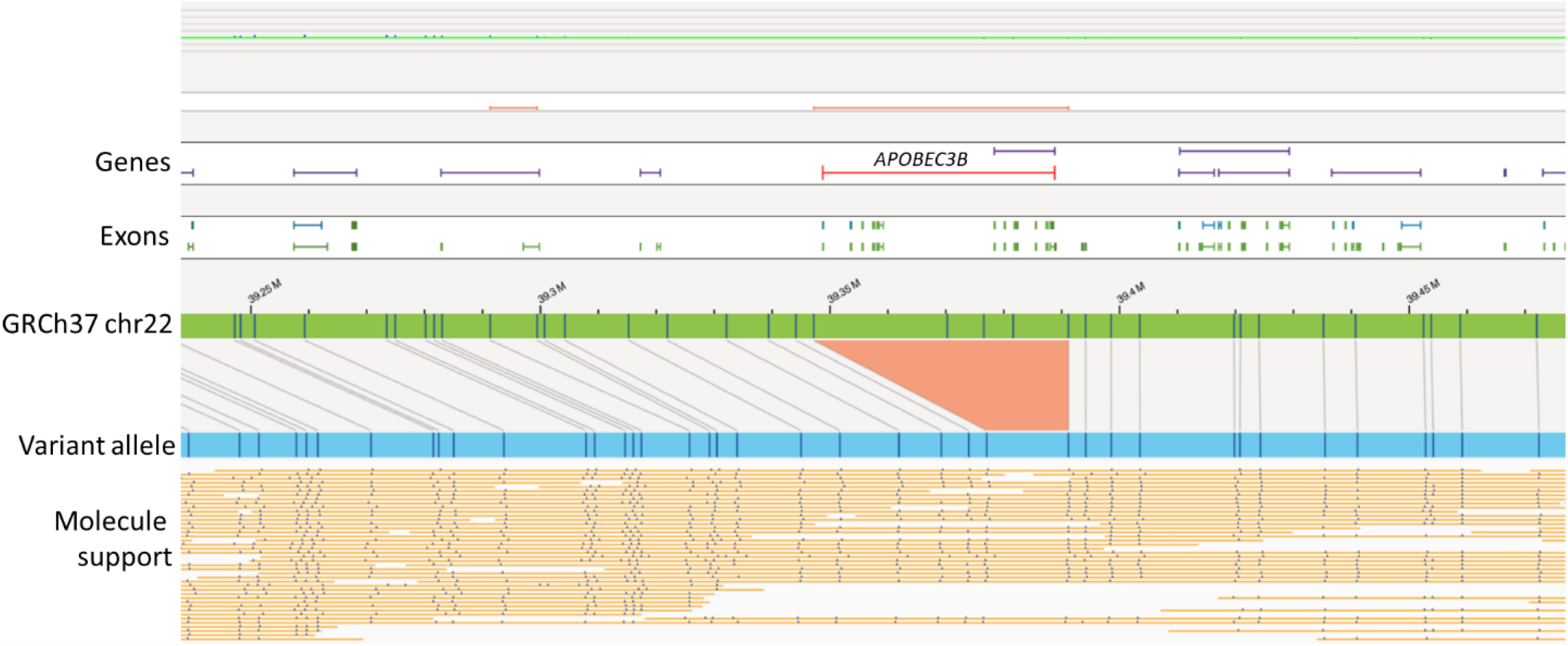
A ∼28.9 kb heterozygous deletion of chr 22 (chr22:39347202-39391164) disrupting 2 genes (*APOBEC3B* and *APOBEC3A*) in three patients.

**Figure 3d:**
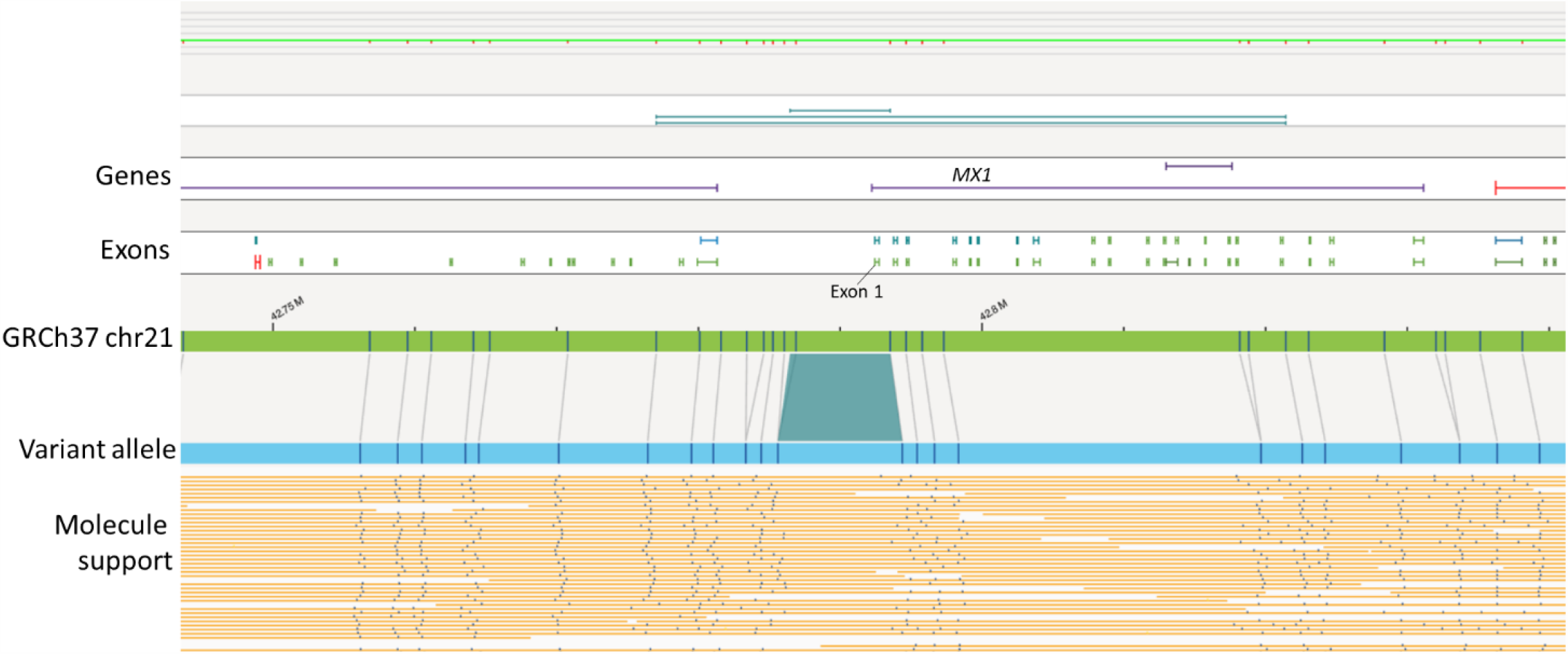
A ∼1.5 kb insertion in chr 21 (chr21:42784603-42793518) overlapping *MX1* gene (exon 1; NM_001144925.2) in 11 patient (cases 13, 17, 18, 20, 21, 24, 25, 28, 4, 8, 38).

### Genome-wide SV enrichment in severely ill vs. population

The odds ratio was calculated comparing SVs identified in the 37 severely ill patients with the population dataset (150 individuals). The data was enriched by directly filtering with numbers of cases on each gene (greater than 2 in the severely ill group) and filtering for SVs that appear in less than 20% frequency in the Bionano controls. The top three gene were found to be *TRPV1, MIR4710*, and *SUMO3*, with a total of 242 genes enriched with log odds ratio greater than three (**Supplementary file 6**).

### Confirmation of Saphyr SVs by FaNDOM and qPCR

FaNDOM confirmed all the 11 SV calls (seven rare/unique + four common) made by Bionano Access Software. Of the 11 SVs, nine events (**Table 2**: all events, **Table 3**: events 2 and 3) were confirmed by qPCR.

### Expression Analysis

To evaluate the impact of X-chromosomal duplication involving the *STK26* gene, *STK26* transcripts were quantified from blood of 11 asymptomatic and 12 severely ill patients with COVID-19, normalized against 18S rRNA. *STK26* transcripts were found to be significantly (7.2 ± 2.3 vs. 1.1 ± 0.5; p<0.001) increased in severely ill patients compared to asymptomatic controls, with the highest expression observed in patient 44 (11.4 fold) (**Figure 4**), who was the one patient that harbored the SV partially duplicating the 5’UTR and coding exon 1 of the *STK26* gene (**Figure 2**).

**Figure 4:**
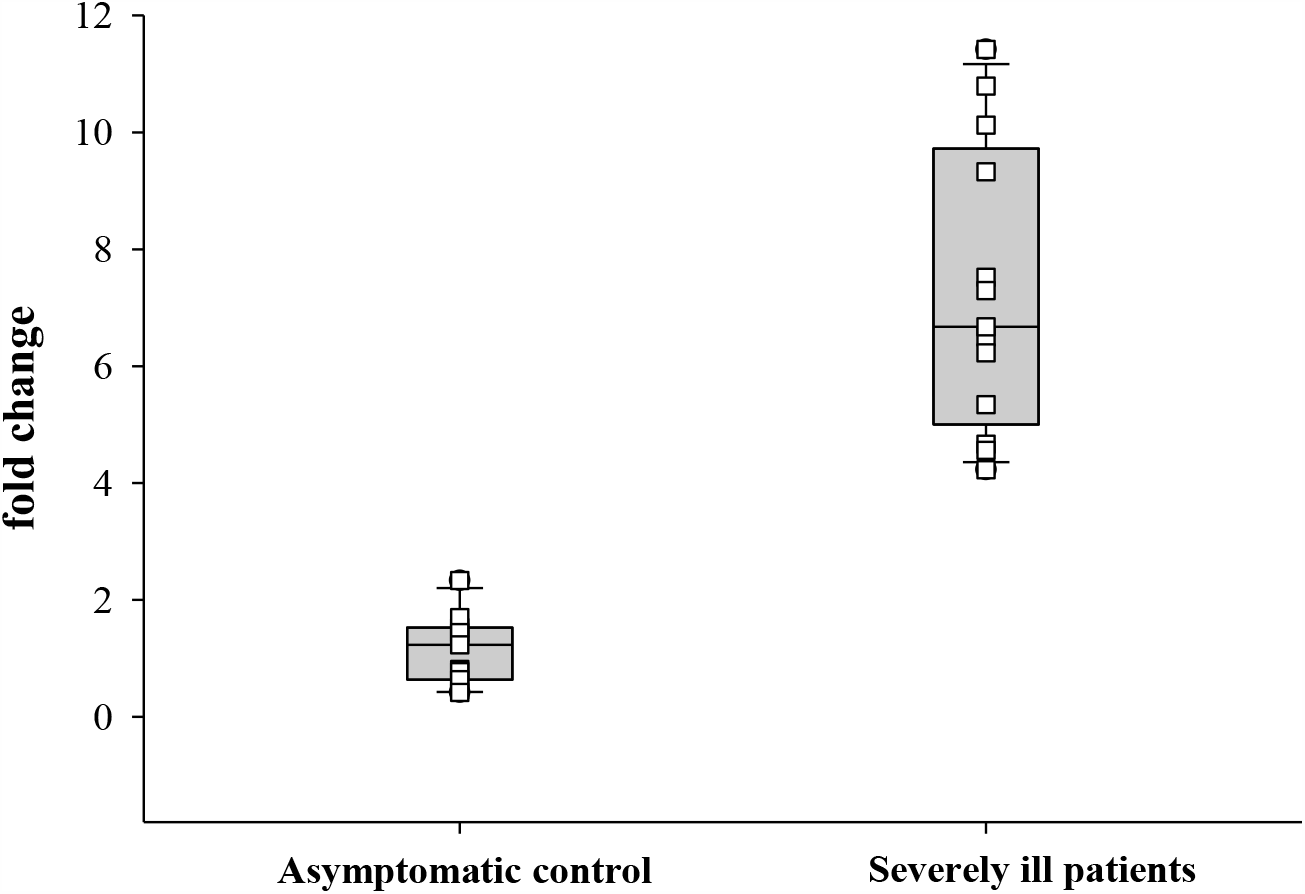
Expression analysis of *STK26*. The *STK26* gene transcripts were quantified in 11 asymptomatic (3 Male and 8 Female) and 12 severely ill patients (6 Male and 6 Female) with COVID-19, normalized against 18S RNA. The *STK26* transcripts were found to be significantly (p<0.001) increased in severely ill patients compared to asymptomatic controls, with the highest expression observed in patient 44, which harbored the SV partially duplicating the *STK26* gene.

## Discussion

This study addressed the current technology bias of short read sequencing that has confined most host genome studies to interrogation of only SNVs and small indels associated with COVID-19 severity [**13-18**]. Although these approaches have been successful in associating certain genetic loci, but large gaps remain in explaining the wide diversity of clinical responses. We aimed to investigate large SVs to identify events that might further explain the inter-individual clinical variability in response to COVID-19, using optical genome mapping. Among 37 patients with severe COVID-19, we found seven rare/private SVs (not found in 267 Bionano controls) potentially impacting 31 genes, and four common SVs impacting seven genes. While individual effect of any of these SVs remains difficult to predict, several of these may be considered candidate loci as strongly predisposing factors associated with severe COVID-19. Altogether, these SVs were implicated in three key host-viral interaction pathways: 1) innate immunity and inflammatory response (*EDARADD, DPP4, ZDHHC1, STK26, MX1, TBC1D3B, CCL3L1, CCL4L2, CD300* gene cluster); 2) airway resistance to pathogens (airway mucous secretion - *MUC4*); and 3) viral replication, spread and RNA editing (*KRT15, KRT19, ZNF443, APOBEC3A, APOBEC3B*) (**Figure 5**).

**Figure 5:**
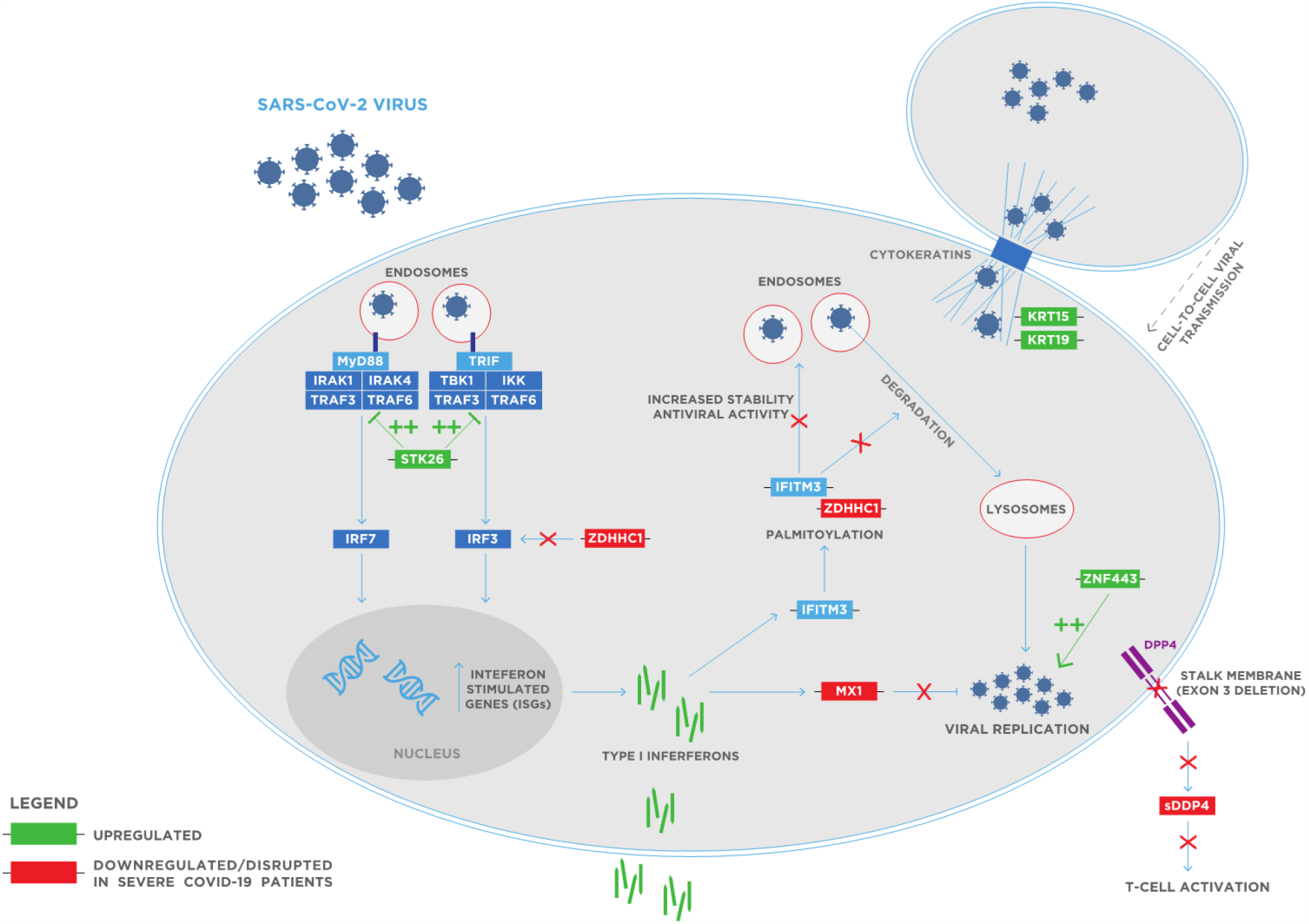
A diagram of the key viral host interaction pathways highlighting genes found to contain structural variation in COVID-19 patients.

### SVs affecting genes implicated in immune and inflammatory response

In this investigation, five rare/unique SVs impacting key immune genes (*STK26, ZDHHC1, DPP4, EDARADD* and *CD300* genes cluster) were identified in six patients. The variability in the host innate and adaptive immune responses to viral infections influence clinical manifestations and outcomes [**34, 35**]. Innate immune response is the first line of defense against the invading pathogen, and utilizes multiple pattern recognition receptors of which, Toll-like receptors (TLRs) have been implicated as key receptors in the recognition of ssRNA of MERS-CoV and SARS-CoV in murine models [**36, 37**]. Independent studies have identified rare genetic variants predicted to be loss of function (LOF) at 13 loci governing TLR3 and TLR7 pathway genes and affecting the type I interferon response [**17, 18]**. In addition, several groups have identified impaired type I interferon response in severe COVID-19 pathogenesis [**19, 38]**. In this study, a rare/unique SV, ∼534.9 kb tandem duplication of chr X was identified in one female patient (case 44). The SV partially disrupts *STK26*, duplicating the 5’UTR and the coding exon 1 region of the gene. In synthetic biology, dual 5’UTR constructs have shown increased transcription and translation of the gene [**39**]. Consistent with this hypothesis, this patient had the highest fold increase of mRNA transcripts as compared to asymptomatic COVID-19 patients, possibly exacerbating the underlying pathogenic mechanism. *STK26* encodes for serine/threonine kinase 26, a negative regulator of the TLR signaling [**40**], directly binds and phosphorylates TRAF6 at two threonine residues (T463 and T486) in the C-terminal TRAF domain and inhibits the oligomerization and autoubiquitination activity of TRAF6 [**41**]. TRAF6 is a downstream signaling molecule in the TLR3/TLR7 pathways that mediates the NF-kB and type I interferon response [**17, 42**]. It is intriguing to speculate that the overexpression of *STK26* in this patient may inhibit the TLR signaling, and via this route impairing the type I interferon response leading to reduced viral clearance in the early phase of the infection and therefore increasing the COVID-19 severity. The upregulation of *STK26* detected coincidentally in all severe COVID-19 patients implicates this gene/pathway in the pathogenesis of severe COVID-19.

The *ZDHHC1* gene deleted in a ∼146.8 kb heterozygous region in one patient (case 39), is also a gene implicated in the type I interferon response. *ZDHHC1* has been shown to mount a type I interferon response against DNA and RNA viruses via different downstream players [**43-45**]. *ZDHHC1* is an endoplasmic reticulum-associated protein that mediates MITA-dependent IRF3 activation and type I interferon response against DNA viruses (**Figure 5**) [**43, 44**]. Predominantly, in RNA virus infection, ZDHHC1 mediates IFITM3 palmitoylation, which ensures the antiviral activity of IFITM3. IFITM3, localized in the endosomal and endolysosomal compartments of cells, prevents the viral entry through the lipid bilayer into the cytoplasm, preventing viral fusion with cholesterol-depleted endosomes against multiple viruses including influenza, SARS-CoV, and HIV-1 [**43-45**]. Knockdown of ZDHHC1 leads to compromised ZDHHC1/IFITM3 anti-viral activity against Japanese encephalitis virus [**45**]. Additionally, IFITM3 expression was found to be upregulated early in epithelial lung cells in response to SARS-CoV-2, [**46**] and that SNPs in IFITM3 are associated with more severe disease [**47**]. We hypothesize that the heterozygous deletion of the *ZDHHC1* gene could possibly compromise the type I interferon response, especially at the level of IFITM3, which might have led to the severe COVID-19 symptoms in this patient.

The rare/unique SV involving *DPP4* gene in a severely ill patient (case 38) is consistent with prior studies implicating DPP4 (protein) in MERS-CoV [**48-50**] and severe COVID-19 [**51**]. *DPP4* exists in two forms: 1) a membrane bound glycoprotein receptor; and 2) a cleaved product of the receptor (sDPP4) that lacks the intracellular and transmembrane domain present in the circulation (**Figure 5**) [**52**]. The ∼24.06 kb heterozygous deletion on chr2 is an intragenic copy number loss of the *DPP4* gene (exon3 and 4). Exons 3 and 4 encode the flexible stalk and α/β hydrolase domain of the receptor. The flexible stalk serves as the cleavage site recognized by matrix metalloproteinase cleavage enzymes [**53**]. The deletion of the two domains is hypothesized to lead to loss of function and significantly low sDPP4 levels. Although bioinformatic models predicted human DPP4-SARS-CoV-2 protein interactions and protein docking studies show SARS-CoV-2 virus with DPP4 as a co-receptor (not as strong as with ACE-2) [**54**], *in vitro* studies found that DPP4 serves as a viral entry receptor for MERS-CoV, and not for coronaviruses including SARS-CoV-2 [**55**]. Apart from the membrane bound receptor, the circulating sDPP4 enzyme interacts with GLP-1 and cytokines and regulates the T-cell receptor mediated T-cell activation. The circulating sDPP4 levels have been found to be reduced in both MERS-CoV and SARS-CoV-2 infection, but to contrasting effects [**50, 51]**. In MERS-CoV infection, the sDPP4 levels were below the threshold needed to exert an antiviral effect, as sufficient viral particles were available for entry via the membrane bound receptor. The reduced sDPP4 levels possibly demonstrate a compromised immune response and does not represent a competition for SARS-CoV-2 entry via the DPP4 receptor. Several case reports and Japanese Adverse Drug Event Report (JADER) has highlighted the risk and incidence of interstitial pneumonia with DPP4 inhibitor (vildagliptin) [**56-58**]. Further, loss of DPP4 activity is associated with a prothrombotic state in myocardial microvessels due to the upregulation of the procoagulant tissue factor [**59**], and reduced post-operative DPP4 activity has been associated with worse patient outcome after cardiac surgery due to paradoxical impairment of angiogenesis and endothelial function [**60, 61**]. Taken together, the partial deletion of *DPP4* in this patient might further contribute to the reduced sDPP4 levels and the compromised immune response predisposing to severe COVID-19.

The rare/*unique* 833 kb **tandem** duplication of 14 genes (*RPL38, MGC16275, TTYH2, Z49982, DNAI2, CD300E, CD300LD, CD300C, CD300LB, CD300A, GPRC5C, GPR142, BTBD17, KIF19*) in one patient (case 19) includes the cluster of *CD300* genes that constitute an important family of receptors expressed on immune cells (myeloid and lymphoid). The CD300 molecules have been identified to participate in mechanisms employed by viruses to escape the immune response and infect host cells [**62, 63**]. Further, CD300 molecules downregulate the cytolytic activity of natural killer (NK) cells against infected cells [**63**]. The duplication of the region involving CD300 genes in this patient may lead to overexpression of these genes that help the virus to evade the immune response leading to disease severity upon infection.

The 162.2 kb duplication partially duplicating the *EDARADD* gene is among the three genes identified to cause ectodermal dysplasia (ED) and is involved in NF-kB activation [**64, 65**]. The two patients with this SV did not have any form of ectodermal dysplasia and the SV was not found in any public database (DGV and gnomAD, last accessed 11/8/2020). To our knowledge, nine *EDARADD* pathogenic sequencing variants have been described, three leading to an autosomal recessive inheritance and six to autosomal dominant mode of inheritance [**66]**. ED caused by variants in the *EDARADD* gene is associated with recurrent pulmonary infections, accompanied by bronchospasm that requires steroidal treatment, and have been found to impair NF-ĸB signaling [**66**]. Cluzeau *et al*., have identified a homozygous deletion of exon 4 in a Tunisian family with severe ED that resulted in complete abolition of NF-KB signaling [**66**]. In addition, several reports have shown that a heterozygous mutation in *EDARADD* in patients with ECTD11A leads to severe impairment of NF-ĸB activation [**67, 68**]. Further, Wohlfart *et al*., reported a heterozygous missense variant in the *EDARADD* gene that did not impact the interaction between EDAR and EDARADD proteins, but led to an impaired ability of the gene to activate NF-ĸB signaling [**68**]. Notably, this patient did not exhibit a phenotype consistent with autosomal dominant ED. Further investigation of the functional consequences of these variants is needed for a more informed interpretation.

In addition to these *rare/private* events, two common SVs impact a cluster of chemokine genes (*TBC1D3B, CCL3L1, CCL4L2*) in 10 of the 37 patients. The CCL3L-CCL4L locus is a copy number variable region (CNVR) with inter-individual variability in breakpoints, and has been extensively associated with HIV-related outcomes [**69**]. A corollary of our findings, single-cell RNA sequencing analysis has found significantly upregulated CCL4L2 in the alveolar macrophages and lung epithelial cells in severely ill compared to mild patients with COVID-19, respectively [**70, 71**]. The duplication of CCL4L2 might explain the upregulation of this chemokine observed in the severely ill patients with COVID-19. Considering chemokine storm to be an important clinical manifestation in severely ill patients, this CNVR region seems to be a critical locus that might influence disease severity by enhancing mobilization and activation of inflammatory cells in these patients. Notably, this SV has a higher frequency in African Americans in our cohort and might be an independent association that could help explain the higher predisposition of the African American population to severe COVID-19. The insertion in the enhancer/promoter region of the *MX1* gene observed in eleven patients could affect regulation of this gene. Several studies have found reduced MX1 expression downstream of a diminished interferon type I and III pathway in patients with severe COVID-19 [**17, 18, 38]**.

While these findings are preliminary in the above-mentioned immune genes, some candidate genes/loci suggests interesting follow-up opportunities. Generally, some observations seem to be in-line with individual variants affecting key immune response genes - collectively they may well explain a few percent of all severe/life-threatening COVID-19 cases (Zhang et al Science, vd Made JAMA). The highlighted rare/ unique SVs all deserve further follow-up before any conclusions can be drawn. These may include segregation analyses in the respective families; and additional downstream expression analysis or protein levels in patient plasma as well as functional studies. It would also be of interest to learn whether any of the identified candidate genes harbor rare/private point mutations (SNVs/InDels) in WES/WGS data of severe COVID-19 cases or show altered expression levels in larger cohorts of severe COVID-19 cases.

### Common SVs disrupting genes of mucociliary axis

The ∼248.1 kb heterozygous inversion disrupting the *MUC4* gene identified in five patients suggests it as a candidate to play a role in increasing disease severity. The mucins have been found to play a critical role in host defense against several respiratory pathogens that include pseudomonas aeruginosa (PA), respiratory syncytial virus (RSV), influenza viruses, and SARS-CoV-2 [**72, 73**]. Respiratory mucous functions as the primary barrier against the inhaled pathogen(s) [**75**]. The epithelium of the respiratory tract is coated with mucous and is the site of primary interaction of the inhaled pathogen with the host. Mucous maintains the hydration of the respiratory tract, traps particulate matter including pathogens, and regulates the immune response [**76**]. Of the 15 mucins in the human respiratory tract, MUC4 is among the five major mucins and belongs to the membrane-tethered class (*MUC1, MUC4*, and *MUC16*) of mucins [**76**]. Tethered mucins play a number of roles, including the activation of intracellular signal transduction pathways, regulation of the immune response, cell differentiation and proliferation [**77**]. In human studies using single-cell RNA sequencing of bronchoalveolar lavage fluid (BALF) cells from patients infected with SARS-CoV-2, upregulation of all the five major mucins was observed [**74**]. Interestingly, the overexpression of *Muc5ac* in mice was protective against influenza virus infection [**78**], whereas, *Muc5b*^-/-^ mice showed a number of severe deficiencies in airway function, reduced mucociliary clearance and severe infections [**79**]. *Muc1*^-/-^ mice have been found to be more susceptible to greater lung injury due to inflammation following PA challenge [**80]**. The anti-inflammatory role of *MUC1* was also observed in an RSV in vitro challenge model [**81**]. Further, female *Muc4*^-/-^ mice develop severe illness that include weight loss, difficulty in breathing, and increased inflammatory cytokines following infection with SARS-CoV-2 [**82**]. Based on the literature evidence and the SVs detected in our cohort, we hypothesize that the disruption of this gene of the mucociliary axis could play a meaningful role in predisposing patients to severe illness following SARS-CoV-2 infection.

### Rare and common SVs affecting genes implicated in viral spread and replication

Two rare/unique SVs impacting the keratin genes (*KRT15* and *KRT19)*, and *ZNF443*, and one common SV impacting *APOBEC3A* and *APOBEC3B* genes were found in six patients. A rare/unique SV, with a ∼ 28.4 kb duplication including two genes (*KRT15* and *KRT19*) was identified in one patient (case 2). The keratins are intermediate filament proteins responsible for the structural integrity of epithelial cells and are part of the cytoskeletal structure of host cells. The keratins, including *KRT15* have been identified to play a major role in the cell-to-cell transmission (i.e. spread) of influenza and SARS-CoV-2 viruses, and have been found to be upregulated in the respiratory tract during infection [**83, 84**]. Another rare/unique duplication was found in the *ZNF443* gene, which has sequence homology to the SARS-CoV-2 virus and might aid in the replication process [**85, 86**]. Duplication of these genes could lead to overexpression and it is hypothesized that this might assist viral replication and spread leading to increased disease severity.

The 28.9 kb heterozygous deletion disrupting *APOBEC3B* and *APOBEC3A* is a known polymorphism that results in a hybrid allele that has the promoter and coding region of *APOBEC3A* but the 3’ UTR of *APOBEC3B*. The apolipoprotein B messenger RNA-editing, enzyme-catalytic, polypeptide-like 3 (APOBEC3) family of cytidine deaminases plays an important role in the innate immune response to viral infections by editing viral genomes [**87, 88**]. The deletion polymorphism has been found to result in decreased expression of both genes [**89**], and is associated with susceptibility to HBV, HIV and HPV infections [**90-92**]. A recent study analyzing the RNA sequences from BALF demonstrated RNA editing marks typical of the ADAR and APOBEC deaminases, suggesting a role of these deaminases in restricting viral propagation [**93**]. The RNA editing deaminasess are critical in determining the fate of both the host and the virus, and the finding of this polymorphism that occurred in three severely ill patients (4.5% in DGV, 11% in gnomAD, and 20% in Bionano controls) warrant further analysis of this key polymorphism which might be related to the variable clinical outcomes in patients with COVID-19.

## Conclusion

The current study provides a powerful framework for investigating structural variations in host genomes that might predispose to severe COVID-19. To our knowledge, this is the first study investigating structural variation using optical genome mapping in host virus interactions in SARS-CoV-2 infected patients. The diverse inter-individual variability observed in the human population after SARS-CoV-2 infection attributed to the disease severity, transmissibility, viral titers, and immune response highlights that numerous pathways/ genes are involved in this complex host-viral interaction. It is insufficient to consider only genomic variation at the level of SNVs/ small indels to understand host variability in response COVID-19 exposure. The present investigation highlights that genes implicated in distinct pathways of host-viral interaction were disrupted by structural variations in patients with severe COVID-19. The events affecting these genes might have predisposed these patients towards severe disease course. Further, several findings in this study are consistent with and support the previous reports implicating the TLR3/TLR7 pathway and type I interferon response in the pathogenesis of COVID-19. One finding, a partial duplication of the TLR negative regulator, *STK26*, and the coincident upregulation of *STK26* is especially intriguing. Incredibly, this finding led us to discover that, relative to levels in asymptomatic COVID-19 cases, *STK26* was upregulated in all 12 tested patients with severe COVID-19. This overexpression may be part of a more widespread severe response profile.

There are some limitations in the present investigation, which will benefit from additional follow-up studies. First, the study is limited to a small patient population, and the findings would be made stronger by validation in a larger cohort. Second, better matched controls for the genomic analysis, such as infected close relatives with distinct clinical manifestations (mild/ asymptomatic), would help control for the confounding influences of common environmental factors. Gaining access to more ideal control samples and performing optical mapping in larger cohorts was beyond the scope of this initial study. Thus, to address these limitations, the following measures were implemented: comparing the frequency of SVs to Bionano control dataset and public databases (DGV and gnomAD), and identifying unique SVs that do not appear in controls and SVs that overlap/ disrupt coding region of the genes implicated in host-viral interaction. Thirdly, functional experiments were not performed to confirm downstream effect of all the SVs in respective genes, because of limited volume of blood available from these patients and no access to their post-mortem lung biopsies. Nevertheless, the whole genome investigation of 37 severely ill patients with COVID-19 identified candidate loci that may provide novel insights into the pathogenesis of severe COVID-19 in some cases.

## Data Availability

All relevant data has been provided in the manuscript and in the supplementary files.

