## Supplementary file 4 for "Host genome analysis of structural variations by Optical Genome Mapping provides clinically valuable insights into genes implicated in critical immune, viral infection, and viral replication pathways in patients with severe COVID-19"

EDARADD_Ex-5

Relative Gene Dosage

|  |  |  |  |  |  |  |  |
| --- | --- | --- | --- | --- | --- | --- | --- |
| Control_M-1 | 0.3 | 0.4 | 0.6 | 0.4 | 0.1 | 0.4 | 1.08654 |
| Control_F-1 | 0.3 | 0.4 | 0.6 | 0.4 | 0.1 | 0.4 | 0.920357 |
| Control_M-1 | 0.3 | 0.4 | 0.6 | 0.4 | 0.1 | 0.4 | 1.08654 |
| Control_F-1 | 0.3 | 0.4 | 0.6 | 0.4 | 0.1 | 0.4 | 0.920357 |
| P-1 | 0.3 | 0.4 | 0.6 | 0.4 | 0.1 | 0.4 | 1.01518 |
| P-22 | 0.3 | 0.4 | 0.6 | 0.4 | 0.1 | 0.4 | 1.79813 |
| P-26 | 0.3 | 0.4 | 0.6 | 0.4 | 0.1 | 0.4 | 1.93529 |

S.4_F-1. qPCR demonstrating a copy number gain of *EDARADD* (exon-5) in patients 22 and 26.

DPP4_Ex-3

Relative Gene Dosage

|  |  |  |  |  |  |  |  |
| --- | --- | --- | --- | --- | --- | --- | --- |
| Control_M-1 | 0.3 | 0.4 | 0.6 | 0.4 | 0.1 | 0.4 | 0.826142 |
| Control_F-1 | 0.3 | 0.4 | 0.6 | 0.4 | 0.1 | 0.4 | 1.02272 |
| Control_M-2 | 0.3 | 0.4 | 0.6 | 0.4 | 0.1 | 0.4 | 1.01297 |
| Control_F-2 | 0.3 | 0.4 | 0.6 | 0.4 | 0.1 | 0.4 | 1.1684 |
| P-38 | 0.3 | 0.4 | 0.6 | 0.4 | 0.1 | 0.4 | 0.508215 |
| P-44 | 0.3 | 0.4 | 0.6 | 0.4 | 0.1 | 0.4 | 0.992633 |
| P-39 | 0.3 | 0.4 | 0.6 | 0.4 | 0.1 | 0.4 | 0.949497 |

S.4_F-2a. qPCR demonstrating a copy number loss of *DPP4* (exon-3) in patient 38.

DPP4_Ex-4

Relative Gene Dosage

|  |  |  |  |  |  |  |  |
| --- | --- | --- | --- | --- | --- | --- | --- |
| Control_M-1 | 0.3 | 0.4 | 0.6 | 0.4 | 0.1 | 0.4 | 0.915449 |
| Control_F-1 | 0.3 | 0.4 | 0.6 | 0.4 | 0.1 | 0.4 | 0.984818 |
| Control_M-2 | 0.3 | 0.4 | 0.6 | 0.4 | 0.1 | 0.4 | 0.959106 |
| Control_F-2 | 0.3 | 0.4 | 0.6 | 0.4 | 0.1 | 0.4 | 1.15649 |
| P-38 | 0.3 | 0.4 | 0.6 | 0.4 | 0.1 | 0.4 | 0.501251 |
| P-44 | 0.3 | 0.4 | 0.6 | 0.4 | 0.1 | 0.4 | 0.996166 |
| P-39 | 0.3 | 0.4 | 0.6 | 0.4 | 0.1 | 0.4 | 0.937122 |

S.4_F-2b. qPCR demonstrating a copy number loss of *DPP4* (exon-4) in patient 38.

ZDHHC1_EX-3

Relative Gene Dosage

|  |  |  |  |  |  |  |  |
| --- | --- | --- | --- | --- | --- | --- | --- |
| Control_M-1 | 0.3 | 0.4 | 0.6 | 0.4 | 0.1 | 0.4 | 1.01792 |
| Control_F-1 | 0.3 | 0.4 | 0.6 | 0.4 | 0.1 | 0.4 | 0.982398 |
| Control_M-1 | 0.3 | 0.4 | 0.6 | 0.4 | 0.1 | 0.4 | 1.01792 |
| Control_F-1 | 0.3 | 0.4 | 0.6 | 0.4 | 0.1 | 0.4 | 0.982398 |
| P-39 | 0.3 | 0.4 | 0.6 | 0.4 | 0.1 | 0.4 | 0.589449 |
| P-38 | 0.3 | 0.4 | 0.6 | 0.4 | 0.1 | 0.4 | 0.88231 |
| P-44 | 0.3 | 0.4 | 0.6 | 0.4 | 0.1 | 0.4 | 0.842409 |

S.4_F-3. qPCR demonstrating a copy number loss of *ZDHHC1* (exon 3) in patient 39.

KRT15-EX-1

Relative Gene Dosage

|  |  |  |  |  |  |  |  |
| --- | --- | --- | --- | --- | --- | --- | --- |
| Control_M-1 | 0.3 | 0.4 | 0.6 | 0.4 | 0.1 | 0.4 | 0.817263 |
| Control_F-1 | 0.3 | 0.4 | 0.6 | 0.4 | 0.1 | 0.4 | 1.05955 |
| Control_M-2 | 0.3 | 0.4 | 0.6 | 0.4 | 0.1 | 0.4 | 1.08991 |
| Control_F-1 | 0.3 | 0.4 | 0.6 | 0.4 | 0.1 | 0.4 | 1.05955 |
| P-2 | 0.3 | 0.4 | 0.6 | 0.4 | 0.1 | 0.4 | 1.67056 |
| P-38 | 0.3 | 0.4 | 0.6 | 0.4 | 0.1 | 0.4 | 0.960382 |
| P-39 | 0.3 | 0.4 | 0.6 | 0.4 | 0.1 | 0.4 | 0.929217 |

S.4_F-4. qPCR demonstrating a copy number gain of *KRT15* (exon-1) in patient 2.

CD300A-EX-2

Relative Gene Dosage

|  |  |  |  |  |  |  |  |
| --- | --- | --- | --- | --- | --- | --- | --- |
| Control_M-1 | 0.3 | 0.4 | 0.6 | 0.4 | 0.1 | 0.4 | 1.11612 |
| Control_F-1 | 0.3 | 0.4 | 0.6 | 0.4 | 0.1 | 0.4 | 1.05885 |
| Control_M-2 | 0.3 | 0.4 | 0.6 | 0.4 | 0.1 | 0.4 | 1.02489 |
| Control_F-2 | 0.3 | 0.4 | 0.6 | 0.4 | 0.1 | 0.4 | 0.825612 |
| P-1 | 0.3 | 0.4 | 0.6 | 0.4 | 0.1 | 0.4 | 1.0695 |
| P-2 | 0.3 | 0.4 | 0.6 | 0.4 | 0.1 | 0.4 | 0.810651 |
| P-3 | 0.3 | 0.4 | 0.6 | 0.4 | 0.1 | 0.4 | 1.03854 |
| P-4 | 0.3 | 0.4 | 0.6 | 0.4 | 0.1 | 0.4 | 0.993121 |
| P-6 | 0.3 | 0.4 | 0.6 | 0.4 | 0.1 | 0.4 | 1.19517 |
| P-7 | 0.3 | 0.4 | 0.6 | 0.4 | 0.1 | 0.4 | 1.01211 |
| P-8 | 0.3 | 0.4 | 0.6 | 0.4 | 0.1 | 0.4 | 1.07488 |
| P-10 | 0.3 | 0.4 | 0.6 | 0.4 | 0.1 | 0.4 | 1.04969 |
| P-11 | 0.3 | 0.4 | 0.6 | 0.4 | 0.1 | 0.4 | 1.20689 |
| P-12 | 0.3 | 0.4 | 0.6 | 0.4 | 0.1 | 0.4 | 1.12177 |
| P-19 | 0.3 | 0.4 | 0.6 | 0.4 | 0.1 | 0.4 | 1.94733 |

S.4_F-5. qPCR demonstrating a copy number gain of *CD300A* (exon-2) in patient 19.

ZNF443-INT-1

Relative Gene Dosage

|  |  |  |  |  |  |  |  |
| --- | --- | --- | --- | --- | --- | --- | --- |
| Control_M-1 | 0.3 | 0.4 | 0.6 | 0.4 | 0.1 | 0.4 | 1.05085 |
| Control_F-1 | 0.3 | 0.4 | 0.6 | 0.4 | 0.1 | 0.4 | 0.975665 |
| Control_M-2 | 0.3 | 0.4 | 0.6 | 0.4 | 0.1 | 0.4 | 0.928151 |
| Control_M-1 | 0.3 | 0.4 | 0.6 | 0.4 | 0.1 | 0.4 | 1.05085 |
| P-1 | 0.3 | 0.4 | 0.6 | 0.4 | 0.1 | 0.4 | 0.93935 |
| P-2 | 0.3 | 0.4 | 0.6 | 0.4 | 0.1 | 0.4 | 0.721229 |
| P-3 | 0.3 | 0.4 | 0.6 | 0.4 | 0.1 | 0.4 | 0.944618 |
| P-4 | 0.3 | 0.4 | 0.6 | 0.4 | 0.1 | 0.4 | 1.17167 |
| P-15 | 0.3 | 0.4 | 0.6 | 0.4 | 0.1 | 0.4 | 0.959977 |
| P-7 | 0.3 | 0.4 | 0.6 | 0.4 | 0.1 | 0.4 | 0.932057 |
| P-19 | 0.3 | 0.4 | 0.6 | 0.4 | 0.1 | 0.4 | 1.21695 |
| P-22 | 0.3 | 0.4 | 0.6 | 0.4 | 0.1 | 0.4 | 1.12669 |
| P-11 | 0.3 | 0.4 | 0.6 | 0.4 | 0.1 | 0.4 | 1.08805 |
| P-12 | 0.3 | 0.4 | 0.6 | 0.4 | 0.1 | 0.4 | 1.07342 |
| P-13 | 0.3 | 0.4 | 0.6 | 0.4 | 0.1 | 0.4 | 1.9279 |

S.4_F-6. qPCR demonstrating a copy number gain of *ZNF443* (intron-1) in patient 13.

STK26_5UTR

Relative Gene Dosage

|  |  |  |  |  |  |  |  |
| --- | --- | --- | --- | --- | --- | --- | --- |
| Control_M-1 | 0.3 | 0.4 | 0.6 | 0.4 | 0.1 | 0.4 | 0.932756 |
| Control_F-1 | 0.3 | 0.4 | 0.6 | 0.4 | 0.1 | 0.4 | 1.08848 |
| Control_F-2 | 0.3 | 0.4 | 0.6 | 0.4 | 0.1 | 0.4 | 1.05595 |
| Control_M-1 | 0.3 | 0.4 | 0.6 | 0.4 | 0.1 | 0.4 | 0.932756 |
| P-44 | 0.3 | 0.4 | 0.6 | 0.4 | 0.1 | 0.4 | 1.34966 |
| P-38 | 0.3 | 0.4 | 0.6 | 0.4 | 0.1 | 0.4 | 1.19802 |
| P-39 | 0.3 | 0.4 | 0.6 | 0.4 | 0.1 | 0.4 | 1.07487 |

S.4_F-7. qPCR demonstrating a copy number gain of *STK26* (5’UTR) in patient 44.
