## Supplementary file 5 for "Host genome analysis of structural variations by Optical Genome Mapping provides clinically valuable insights into genes implicated in critical immune, viral infection, and viral replication pathways in patients with severe COVID-19"

CCL4L2

Relative Gene Dosage

|  |  |  |  |  |  |  |  |
| --- | --- | --- | --- | --- | --- | --- | --- |
| Control_M-1 | 0.3 | 0.4 | 0.6 | 0.4 | 0.1 | 0.4 | 1.0302 |
| Control_F-1 | 0.3 | 0.4 | 0.6 | 0.4 | 0.1 | 0.4 | 0.974585 |
| Control_M-2 | 0.3 | 0.4 | 0.6 | 0.4 | 0.1 | 0.4 | 1.02197 |
| Control_F-1 | 0.3 | 0.4 | 0.6 | 0.4 | 0.1 | 0.4 | 0.974585 |
| P-1 | 0.3 | 0.4 | 0.6 | 0.4 | 0.1 | 0.4 | 0.953011 |
| P-2 | 0.3 | 0.4 | 0.6 | 0.4 | 0.1 | 0.4 | 0.97533 |
| P-3 | 0.3 | 0.4 | 0.6 | 0.4 | 0.1 | 0.4 | 1.15073 |
| P-4 | 0.3 | 0.4 | 0.6 | 0.4 | 0.1 | 0.4 | 0.943022 |
| P-6 | 0.3 | 0.4 | 0.6 | 0.4 | 0.1 | 0.4 | 1.87922 |
| P-8 | 0.3 | 0.4 | 0.6 | 0.4 | 0.1 | 0.4 | 1.47757 |
| P-12 | 0.3 | 0.4 | 0.6 | 0.4 | 0.1 | 0.4 | 1.34383 |
| P-13 | 0.3 | 0.4 | 0.6 | 0.4 | 0.1 | 0.4 | 2.46868 |
| P-19 | 0.3 | 0.4 | 0.6 | 0.4 | 0.1 | 0.4 | 1.70076 |
| P-20 | 0.3 | 0.4 | 0.6 | 0.4 | 0.1 | 0.4 | 1.83081 |
| 0 | 0.3 | 0.4 | 0.6 | 0.4 | 0.1 | 0.4 | 0.00903514 |

CCL4L2

Relative Gene Dosage

|  |  |  |  |  |  |  |  |
| --- | --- | --- | --- | --- | --- | --- | --- |
| Control_M-1 | 0.3 | 0.4 | 0.6 | 0.4 | 0.1 | 0.4 | 1.0302 |
| Control_F-1 | 0.3 | 0.4 | 0.6 | 0.4 | 0.1 | 0.4 | 0.974585 |
| Control_M-2 | 0.3 | 0.4 | 0.6 | 0.4 | 0.1 | 0.4 | 1.02197 |
| Control_F-1 | 0.3 | 0.4 | 0.6 | 0.4 | 0.1 | 0.4 | 0.974585 |
| P-23 | 0.3 | 0.4 | 0.6 | 0.4 | 0.1 | 0.4 | 1.2342 |
| P-25 | 0.3 | 0.4 | 0.6 | 0.4 | 0.1 | 0.4 | 1.21357 |
| P-26 | 0.3 | 0.4 | 0.6 | 0.4 | 0.1 | 0.4 | 1.03734 |
| P-27 | 0.3 | 0.4 | 0.6 | 0.4 | 0.1 | 0.4 | 1.19393 |
| P-21 | 0.3 | 0.4 | 0.6 | 0.4 | 0.1 | 0.4 | 1.44577 |
| P-22 | 0.3 | 0.4 | 0.6 | 0.4 | 0.1 | 0.4 | 3.32476 |
| P-28 | 0.3 | 0.4 | 0.6 | 0.4 | 0.1 | 0.4 | 2.41996 |
| P-29 | 0.3 | 0.4 | 0.6 | 0.4 | 0.1 | 0.4 | 1.88978 |
| 0 | 0.3 | 0.4 | 0.6 | 0.4 | 0.1 | 0.4 | 0.00903514 |
| 0 | 0.3 | 0.4 | 0.6 | 0.4 | 0.1 | 0.4 | 0.00903514 |
| 0 | 0.3 | 0.4 | 0.6 | 0.4 | 0.1 | 0.4 | 0.00903514 |

S.5_F-1. qPCR demonstrating a copy number gain of *CCL4L2* (intron 1) in patients 12, 20, 28, 29, 6, 8 13,19,21,and 22.

APOBEC3A-Ex-1

Relative Gene Dosage

|  |  |  |  |  |  |  |  |
| --- | --- | --- | --- | --- | --- | --- | --- |
| Control_M-1 | 0.3 | 0.4 | 0.6 | 0.4 | 0.1 | 0.4 | 1.10348 |
| Control_F-1 | 0.3 | 0.4 | 0.6 | 0.4 | 0.1 | 0.4 | 1.03448 |
| Control_M-2 | 0.3 | 0.4 | 0.6 | 0.4 | 0.1 | 0.4 | 1.04037 |
| Control_F-2 | 0.3 | 0.4 | 0.6 | 0.4 | 0.1 | 0.4 | 0.842029 |
| P-1 | 0.3 | 0.4 | 0.6 | 0.4 | 0.1 | 0.4 | 1.09892 |
| P-2 | 0.3 | 0.4 | 0.6 | 0.4 | 0.1 | 0.4 | 0.808051 |
| P-3 | 0.3 | 0.4 | 0.6 | 0.4 | 0.1 | 0.4 | 1.08736 |
| P-4 | 0.3 | 0.4 | 0.6 | 0.4 | 0.1 | 0.4 | 1.00429 |
| P-6 | 0.3 | 0.4 | 0.6 | 0.4 | 0.1 | 0.4 | 1.22711 |
| P-7 | 0.3 | 0.4 | 0.6 | 0.4 | 0.1 | 0.4 | 1.08467 |
| P-8 | 0.3 | 0.4 | 0.6 | 0.4 | 0.1 | 0.4 | 1.14237 |
| P-10 | 0.3 | 0.4 | 0.6 | 0.4 | 0.1 | 0.4 | 1.07796 |
| P-11 | 0.3 | 0.4 | 0.6 | 0.4 | 0.1 | 0.4 | 1.22946 |
| P-15 | 0.3 | 0.4 | 0.6 | 0.4 | 0.1 | 0.4 | 0.591601 |
| P-29 | 0.3 | 0.4 | 0.6 | 0.4 | 0.1 | 0.4 | 0.642962 |

S.5_F-2. qPCR demonstrating a copy number loss of *APOBEC3A* (exon 1) in patients 15 and 29.
